# Genetic architecture of the human brain glycoproteome implicates glycosylation in Alzheimer’s disease and related dementias

**DOI:** 10.64898/2026.11.24.26368888

**Authors:** Ru Feng, Anqi Wang, Xuewei Cao, Sheema Sameen, David Morgenstern, Hila Levy, Aparajita Pranjal, Ruixi Li, Haochen Sun, Yining Li, Kushal K. Dey, Nicholas T. Seyfried, David A. Bennett, Philip L. De Jager, The Alzheimer’s Disease Functional Genomics Consortium, Gao Wang, Yishai Levin, Michal Schnaider Beeri, Aron S. Buchman, Hans-Ulrich Klein

**Affiliations:** Center for Statistical Genetics, The Gertrude H. Sergievsky Center, Columbia University, New York, NY, USA; Center for Translational & Computational Neuroimmunology, Department of Neurology, Columbia University Irving Medical Center, New York, NY, USA; Taub Institute for Research on Alzheimer’s disease and the Aging brain, Columbia University Irving Medical Center, New York, NY, USA; Computational and Systems Biology Program, Memorial Sloan Kettering Cancer Center, New York, NY, USA; Nancy and Stephen Grand Israel National Center for Personalized Medicine, Weizmann Institute of Science, Rehovot, Israel; Department of Neurology, Columbia University, New York, NY, USA; Department of Biochemistry, Emory University, Atlanta, GA, USA; Rush Alzheimer’s Disease Center, Rush University Medical Center, Chicago, IL, USA; The Krieger Klein Alzheimer’s Research Center, Brain Health Institute, Rutgers University, NJ, USA

## Abstract

Glycosylation is a prevalent post-translational modification in the human brain, but its genetic regulation remains poorly defined. We profiled N-linked glycosylation in 316 human prefrontal cortices and generated the first glycoproteome-wide map of glycosylation quantitative trait loci (glycoQTLs). GlycoQTLs were enriched in regulatory regions and partly independent of protein abundance. High-mannose glycopeptiforms were preferentially affected, suggesting genetic influence on early glycan processing in the endoplasmic reticulum. Integration with Alzheimer’s disease and related dementias (ADRD) GWAS summary statistics identified disease-associated glycoQTLs, including those at TMEM106B and ACE. At TMEM106B, the ADRD risk allele altered the glycan composition at N183 within the fibril-forming C-terminal fragment of the protein, whereas at ACE, individual glycopeptiforms showed divergent genetic effects. Targeted trans-glycoQTL analysis further identified long-range regulation, including a hotspot at the fucosidase FUCA2. These findings establish glycoQTL mapping as a complementary functional-genomics layer that reveals disease-associated molecular effects beyond conventional protein-level analyses.

## Introduction

Late-onset Alzheimer’s disease (AD) is a complex disease whose genetic contributions have been extensively characterized through genome-wide association studies (GWAS)^1–6^. The downstream effects of AD risk variants on molecular traits in disease-relevant tissues and, more recently, in specific cell types have been mapped through large expression and protein quantitative trait locus (eQTL and pQTL) studies^7–10^. These resources have substantially advanced our understanding of Alzheimer’s disease and related dementias (ADRD). However, in contrast to the transcriptome and proteome, the genetic regulation of post-translational modifications (PTMs) remains poorly defined, and it is not yet clear to what extent ADRD risk variants exert their effects by altering PTMs.

Glycosylation, the enzymatic attachment of glycans to proteins, is one of the most prevalent and structurally diverse PTMs in humans. This diversity arises from the large repertoire of glycan compositions and branching structures, which critically modulate protein folding, stability, trafficking, and function. Glycoproteins are involved in numerous biological processes, including immune signaling, protein quality control, and synaptic communication^11,12^. Aberrant glycosylation has been implicated in several neurological diseases^13^, including AD^14–17^, where glycosylation of β-amyloid-related proteins (APP, BACE1, and ADAM10) may play a role in β-amyloid homeostasis^18–20^. Moreover, glycosylation patterns in cerebrospinal fluid and plasma have been explored as potential AD biomarkers^21,22^.

Although previous studies have demonstrated that common genetic variants can influence glycosylation, these efforts have important limitations^23,24^. Most studies have focused on the glycome, that is, the overall abundance of glycans within a sample, without considering the specific proteins or glycosylation sites on which these glycans reside^25–28^. Such approaches primarily identify genetic regulators of the glycosylation machinery but cannot resolve protein- or site-specific alterations. To date, protein-specific glycosylation QTLs (glycoQTLs) have been reported for only a small number of glycoproteins, largely through targeted studies of immunoglobulin G and transferrin^29–33^, and glycoproteome-wide QTL analyses remain lacking. Moreover, existing glycoQTL studies have been conducted almost exclusively in blood plasma. Substantial evidence suggests that the brain glycoproteome is highly distinct^34–37^, and thus current resources provide limited insight into the role of glycosylation in neurological disease.

In this study, we profiled the glycoproteome of 316 human prefrontal cortex samples from the Religious Orders Study (ROS) and the Rush Memory and Aging Project (MAP)^38^. By integrating these data with matched brain proteomic and whole genome sequencing data from the same individuals, we generated the first large-scale map of glycoQTLs in the human cortex alongside matched pQTLs **(Fig. 1a)**. We then integrated these QTLs with ADRD GWAS summary statistics and investigated selected findings at TMEM106B and ACE in greater detail. Finally, we performed a targeted trans-glycoQTL analysis.

**Figure 1.**
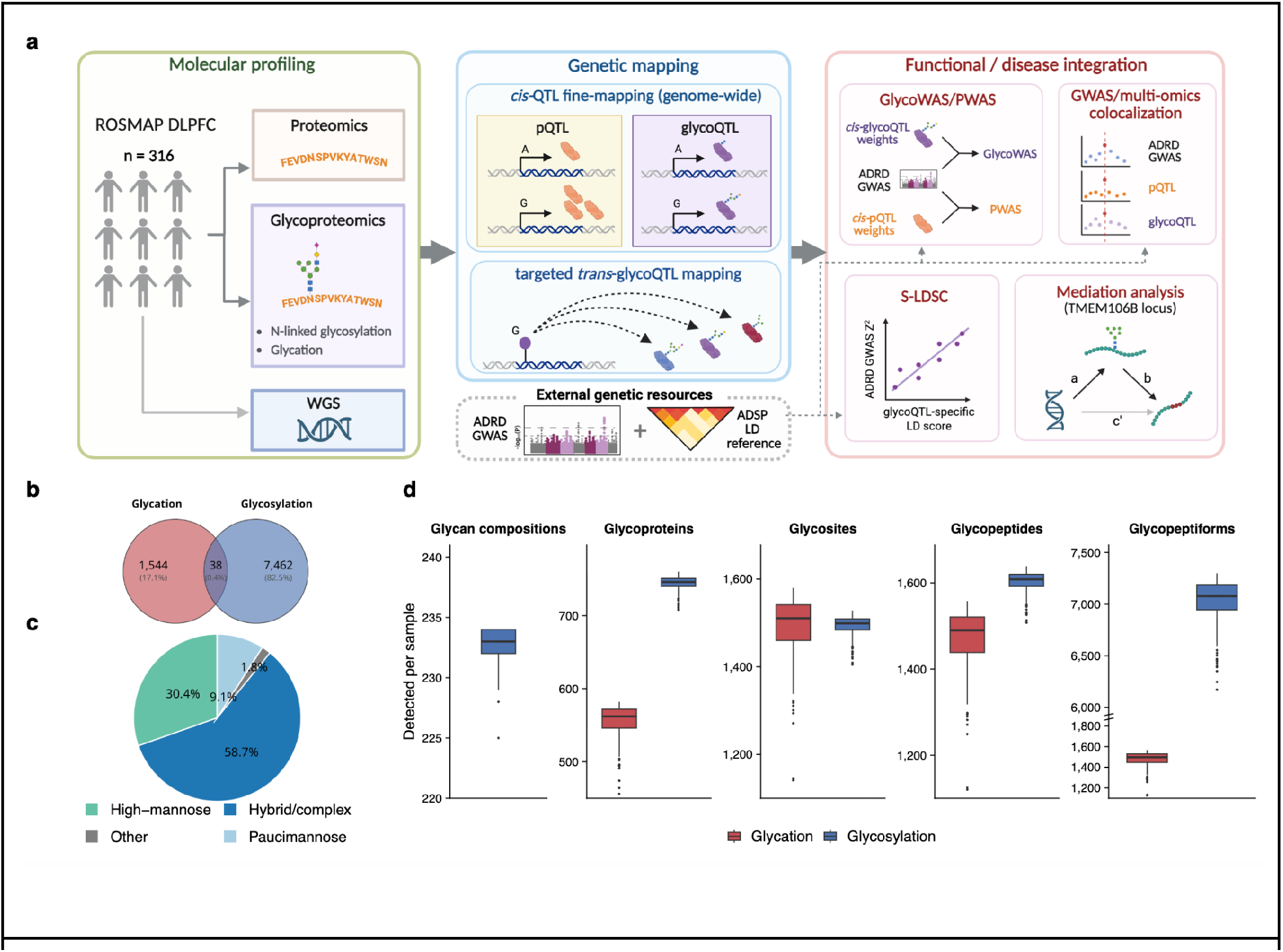
Study design and characterization of the human brain glycoproteome. a) Overview of the experimental design. Protein abundance and glycosylation (glycation and glycosylation) were quantified in DLPFC tissue from the ROSMAP cohort (n=316) alongside whole-genome sequencing (WGS) genotype data. QTL fine-mapping was performed to identify protein QTLs (pQTL) and glycoQTLs (glycoQTL). These QTL results, together with AD GWAS summary statistics and ADSP LD reference data, served as inputs for downstream integrative analyses, including mediation analysis to assess direct and indirect effects, TWAS to link genes and traits, stratified LD score regression (S-LDSC) for heritability enrichment, and colocalization analysis to evaluate shared genetic signals. b) Venn diagram showing the numbers of detected glycated and N-glycosylated glycopeptiforms. The intersection represents glycopeptiforms carrying both glycation and an N-linked glycan. c) Relative abundance of glycan classes among detected glycopeptiforms. d) Number of detected glycan compositions, glycoproteins, glycosites, glycopeptides, and glycopeptiforms per sample. Boxplots summarize the distribution across samples.

## Results

### Comprehensive profiling of the glycoproteome in the aged human brain

The glycoproteome of 316 dorsolateral prefrontal cortex (DLPFC) samples from the ROSMAP cohort **(Table S1)** was profiled using an improved glycopeptide-enrichment workflow followed by liquid chromatography-tandem mass spectrometry (LC-MS/MS)^39^. This approach quantified 9,044 glycopeptiforms, defined as unique peptide-glycan combinations, with a low missing rate of 6.3% across all glycopeptiforms and samples. Most glycopeptiforms (82.5%) carried N-linked glycans, while 17.1% represented peptides with a lysine-linked hexose adduct **(Fig. 1b)**. Unlike glycosylation, this modification reflects a non-enzymatic glycation process that occurs sporadically and is typically associated with elevated glucose levels. The remaining 0.4% of glycopeptiforms contained both an N-linked glycan and a glycated lysine.

The 9,044 detected glycopeptiforms originated from 1,217 distinct glycoproteins, spanning 1,487 N-linked glycosylation sites (glycosites) and 1,564 glycated lysines **(Fig. S1a-b)**. Across all glycosites, we detected 234 compositionally distinct N-glycans. Classification of the N-glycans into four categories, complex/hybrid, high-mannose, paucimannose and other, revealed that high-mannose glycans accounted for a substantial fraction (30.4%) of total glycan abundance **(Fig. 1c),** consistent with previous observations in mammalian brain tissue^35,12,37^. The number of glycopeptiforms per glycoprotein exhibited a strongly right-skewed distribution **(Fig. S1c)**, with a median of 3 and a maximum of 231 when considering N-linked glycosylation only. A similarly skewed distribution was observed for the number of glycosites per glycoprotein **(Fig. S1d),** driven by a small set of highly glycosylated proteins. Detection rates across samples were high, with a median of 7,076.5 glycopeptiforms per sample (IQR: 6,943-7,188) for N-glycosylation and 1,497.5 per sample (IQR: 1,445-1,529) for glycation **(Fig. 1d).** Together, the completeness and the depth of glycopeptiform coverage make this dataset a unique and comprehensive resource for studying the glycoproteome of the aged human brain.

### Genomic fine-mapping identifies widespread glycoQTLs with effects partially independent of protein abundance

To map the genetic regulation of the brain glycoproteome, we performed genome-wide cis-glycoQTL detection and fine-mapping using SuSiE^40^, a Bayesian regression framework that jointly models all variants within a locus while accounting for linkage disequilibrium and allowing for multiple independent effects. Each 95% credible set was considered an independent glycoQTL signal. Using a minor allele frequency (MAF) filter of 5%, this approach identified 206 glycopeptiforms that were regulated by 239 independent QTLs **(Fig. 2a).** Similar results were obtained using conventional marginal QTL mapping with TensorQTL^41^ **(Fig. S2a)**, indicating that parts of the glycoproteome are influenced by genetic variation.

**Figure 2.**
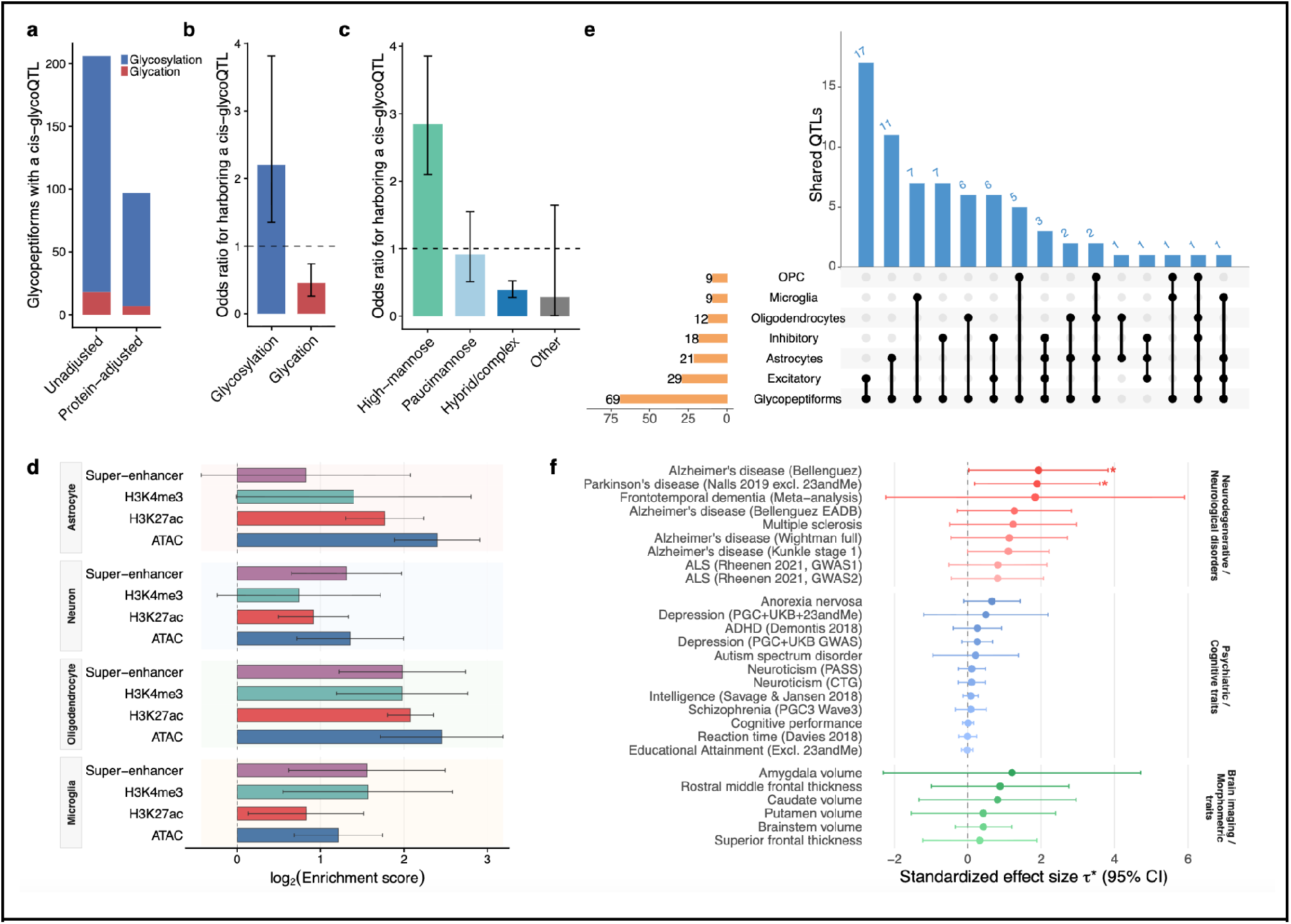
Genetic architecture and functional characterization of glycoQTLs. a) Number of glycopeptiforms with a cis-glycoQTL, summarized separately for glycosylation and glycation under unadjusted and protein-adjusted models. b) Odds ratios for harboring a cis-glycoQTL for glycosylation compared with glycation, and vice versa. Error bars indicate 95% confidence intervals. c) Odds ratios for harboring a cis-glycoQTL across N-glycan classes. For each glycan class, the odds ratio was calculated relative to all remaining glycan classes combined. Error bars indicate 95% confidence intervals. d) Enrichment of fine-mapped cis-glycoQTL variants in cell type-specific ATAC, H3K27ac, H3K4me3, and super-enhancer annotations across four major brain cell types. Error bars indicate 95% confidence intervals. e) Multi-context colocalization of glycoQTLs with cell type-specific eQTLs from six major brain cell types. Bars show the number of shared QTLs for each combination of cell types. (OPC = oligodendrocyte precursor cells) f) Standardized effect sizes (τ*) estimated by stratified LD score regression for glycoQTL annotations across neurodegenerative and neurological disorders, psychiatric and cognitive traits, and brain imaging phenotypes. Error bars indicate 95% confidence intervals.

We next asked to what extent these glycoQTLs reflect genetic regulation of total protein abundance rather than glycosylation-specific effects. Since altered glycopeptiform levels may simply reflect altered total protein levels, we repeated the analysis after linear adjustment of glycopeptiform levels for matched protein abundance. This resulted in a 52.9% reduction in glycopeptiforms with detectable glycoQTLs, suggesting that a considerable fraction of glycoQTL effects may be mediated through protein abundance **(Fig. 2a).** Consistently, colocalization analysis showed that 43.0% of glycoQTLs colocalize with pQTLs **(Fig. S2b).** However, a substantial subset of glycoQTLs persisted after adjustment, indicating that these glycoQTLs reflect genetic effects on glycosylation that are at least partially independent of steady-state protein abundance.

Further evidence for glycosylation-specific effects comes from comparing glycosylated to glycated peptiforms. Since glycation, in contrast to tightly regulated glycosylation, occurs largely through spontaneous non-enzymatic reactions, it is expected to depend primarily on protein abundance. Interestingly, glycosylated peptiforms were more likely (odds ratio = 2.20, P = 7.10 ×10^−4^) to harbor a QTL than glycated peptiforms **(Fig. 2b),** supporting the presence of genetic variants that specifically affect glycosylation. We next examined whether genetic effects differ across glycan classes. N-linked glycosylation is initiated in the endoplasmic reticulum (ER), where high-mannose glycans are generated during protein synthesis, and continues in the Golgi apparatus, where further processing can generate hybrid and complex structures. Interestingly, high-mannose glycopeptiforms were more likely to harbor QTLs than non-high-mannose glycopeptiforms (odds ratio = 2.84, P = 2.91 ×10^−12^), with strong enrichment relative to hybrid/complex glycopeptiforms (odds ratio = 3.02, P = 1.85 ×10^−12^) and a somewhat weaker enrichment relative to paucimannose glycopeptiforms (odds ratio = 1.97, P = 0.0103) **(Fig. 2c).** These results suggest that genetic variants preferentially influence glycosylation states established in the ER.

Lastly, we evaluated the functional context of glycoQTLs across major brain cell types. GlycoQTL variants were enriched in regulatory regions and open chromatin across all major cell types **(Fig. 2d)**, similar to patterns observed for eQTLs and pQTLs^42^. Together, these results suggest that a large number of glycoQTLs could be mediated through gene regulatory mechanisms. One possible explanation is that cis-acting genetic variation influences the timing or rate of protein production and trafficking through the secretory pathway, thereby affecting protein exposure to glycosylation enzymes in the ER and Golgi. Such effects could modulate glycoproteoform composition, particularly early high-mannose states generated during protein synthesis and processing in the ER, even in the absence of detectable changes in steady-state protein abundance.

### Glycosylation QTLs are supported across brain cell types and enriched for neurological disease traits

To assess which cell types contribute to the glycoQTL signals observed at the tissue level, we utilized single-nucleus eQTLs derived from six major brain cell types: excitatory neurons, inhibitory neurons, astrocytes, microglia, oligodendrocytes and oligodendrocyte precursor cells. Since many glycoQTLs colocalize with pQTLs and are enriched in regulatory regions, we hypothesized that glycoQTLs may also colocalize with cell type-specific eQTLs. Multi-context colocalization analysis^43^ showed that glycoQTLs most frequently colocalize with eQTLs in excitatory neurons (29 shared QTLs), followed by astrocytes (21 shared QTLs) **(Fig. 2e)**. However, excitatory neurons also exhibited the highest average number of detectable genes in the snRNA-seq data, suggesting that colocalization frequency largely reflects underlying cell type abundance and eQTL discovery power rather than cell type-specific enrichment **(Sup Fig 2c).** Consistent with this observation, glycoQTLs were enriched in cell type-specific regulatory regions across all major brain cell types **(Fig. 2d).** In summary, these findings suggest that glycoQTL signals detected at the tissue level are represented across all major brain cell types, with more abundant cell populations contributing proportionally more glycoQTL signals.

To explore the potential involvement of glycosylation in complex traits, we applied stratified LD score regression to evaluate the contribution of glycoQTL annotations to per-SNP heritability across neurodegenerative and neurological disorders, psychiatric and cognitive traits, and brain imaging phenotypes **(Fig. 2f).** Glycated peptiforms were excluded from this analysis. Although no trait remained significant after multiple testing correction, several traits showed a nominally significant positive contribution of the glycoQTL annotation to per-SNP heritability and were considered candidates for further investigation. Among neurological diseases, the strongest contribution was observed for Parkinson’s disease^44^ (P = 0.030) and Alzheimer’s disease^3^ (P = 0.046). Given these results, we prioritized Alzheimer’s disease for subsequent integrative analyses of glycoQTLs and GWAS summary statistics.

### Genetically regulated glycopeptiform abundance is associated with Alzheimer’s disease and related dementia risk

To investigate whether genetically regulated variation in the brain glycoproteome is associated with ADRD risk, we integrated glycoQTLs with summary statistics from a recent ADRD GWAS3 using a transcriptome-wide association study framework (TWAS)^45^ adapted for glycopeptiform abundance (glycoproteome-wide association study, GlycoWAS). Among 1,101 glycopeptiforms with sufficient genetic predictability, GlycoWAS identified 42 significant glycopeptiforms from 17 glycoproteins after Bonferroni correction (P < 4.54 ×10^−5^) **(Fig. 3a; Table S2).** Notably, GlycoWAS performed on glycopeptiform abundances adjusted for matched protein abundance showed attenuated associations, but significant glycopeptiform associations remained for 11 of the 17 glycoproteins **(Fig. S3a),** while a conventional proteome-wide association study (PWAS) identified significant associations for 6 of the 17 glycoproteins **(Fig. S3b).** These findings are consistent with our earlier observation that while many glycoQTLs colocalize with pQTLs, glycoQTLs may alter glycoproteoform composition beyond their effects on total protein abundance.

**Figure 3.**
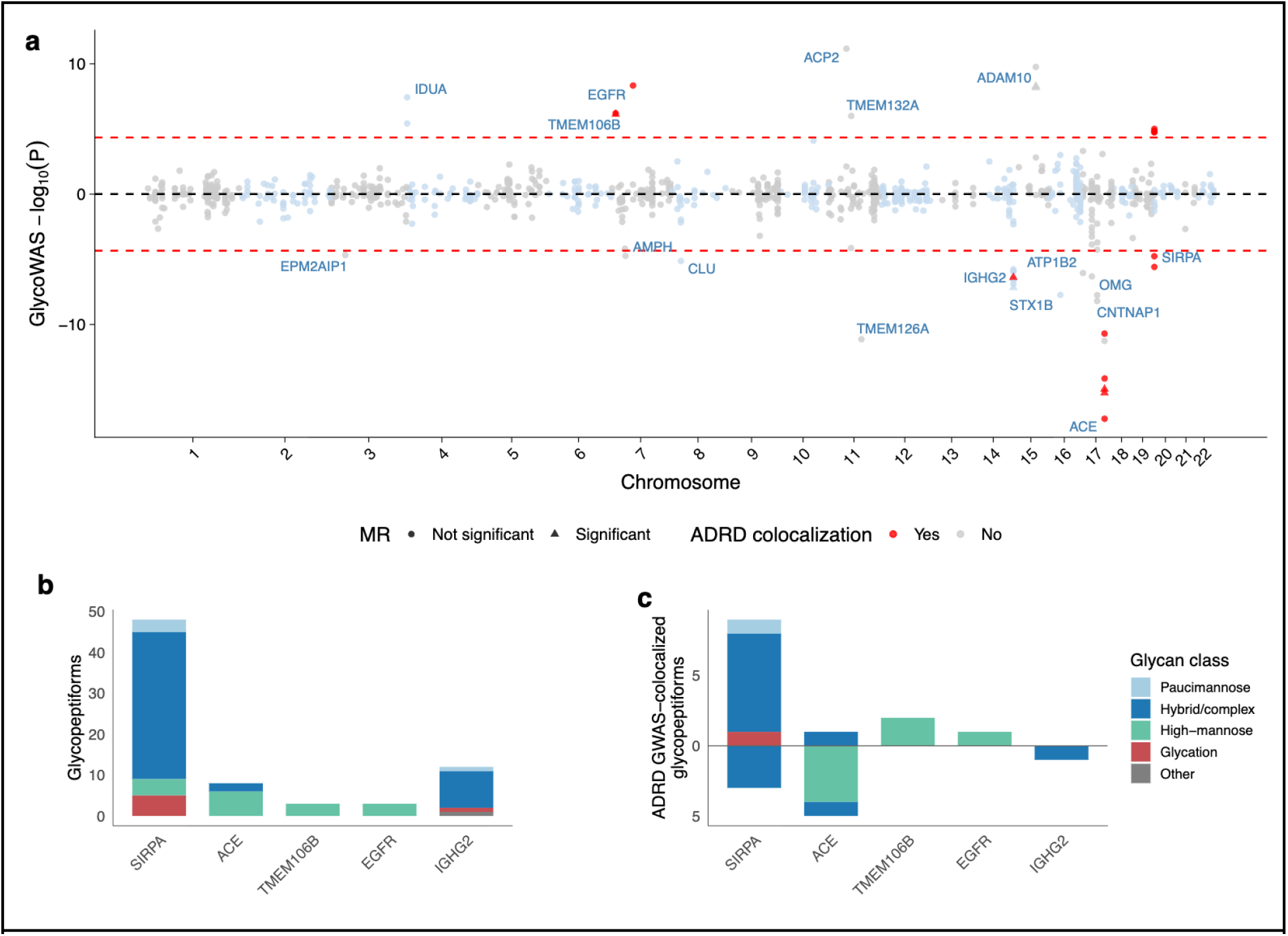
Glycoproteome-wide association analysis of Alzheimer’s disease and related dementia. a) GlycoWAS results using unadjusted glycoproteomic data. Each point represents a glycopeptiform. The association statistic is plotted as the signed log-transformed P value, with positive values indicating that the ADRD risk allele is associated with higher glycopeptiform abundance and negative values indicating lower abundance. Red dashed lines denote the Bonferroni significance threshold. Significant glycopeptiforms are labeled by their corresponding glycoprotein. Among glycopeptiforms with significant GlycoWAS associations, red points indicate glycoQTL colocalization with an ADRD GWAS signal, light blue or gray points indicate no evidence of colocalization, and triangles indicate additional support from Mendelian randomization analysis. b) Total number of analyzed glycopeptiforms for each glycoprotein with at least one glycoQTL colocalizing with an ADRD GWAS signal. Colors indicate glycan class. c) Number of glycopeptiforms with glycoQTLs colocalizing with ADRD GWAS signals for each glycoprotein shown in b, colored by glycan class and separated by glycoQTL effect direction (above zero, ADRD risk allele associated with higher glycopeptiform abundance; below zero, lower glycopeptiform abundance)

To further prioritize loci supported by shared genetic mechanisms, we performed colocalization^43^ analyses integrating glycoQTL and ADRD GWAS signals. Five of the 17 glycoproteins (TMEM106B, EGFR, IGHG2, ACE and SIRPA) showed evidence of colocalization with ADRD risk. These proteins have previously been implicated in biological processes relevant to neurodegeneration, including lysosomal function (TMEM106B), immune signaling (SIRPA and IGHG2), and growth factor or proteolytic pathways (EGFR and ACE). Notably, each of these glycoproteins was represented by multiple detected glycopeptiforms, ranging from 3 to 48 glycopeptiforms per protein (Fig. 3b). However, only a subset of these glycopeptiforms showed glycoQTL effects colocalizing with ADRD GWAS signals (Fig. 3c), further supporting the notion that ADRD risk variants do not uniformly affect all glycoproteoforms of a protein. Intriguingly, in two cases (ACE and SIRPA), distinct glycopeptiforms derived from the same protein showed opposite directions of association with ADRD risk (Fig. 3c). These findings suggest that ADRD risk variants may selectively alter glycoproteoform composition, highlighting molecular effects that are not resolved in a conventional protein-level analysis. In the following sections, we examine two particularly interesting loci, TMEM106B and ACE, in detail.

### TMEM106B risk variant increases C-terminal peptide abundance and preferentially enriches HexNAc(2)Hex(6) glycopeptiforms at N183

TMEM106B is a lysosomal transmembrane protein with five predicted N-glycosylation sites in its luminal C-terminal domain^46^. Higher TMEM106B protein abundance has been reported in carriers of the ADRD risk allele^47^. The candidate coding variant rs3173615 (T185S) lies two residues downstream of the N183 glycosylation site **(Fig. 4a)**, with the protective S185 allele converting the local glycosylation sequon from N-I-T to N-I-S. Although both motifs support N-linked glycosylation, N-X-S sequons are generally glycosylated less efficiently than N-X-T sequons^48^, raising the possibility that the risk variant acts through altered N183 glycosylation. However, the glycoforms present at N183 and whether T185S affects site occupancy, glycan composition, or both remain incompletely characterized.

**Figure 4.**
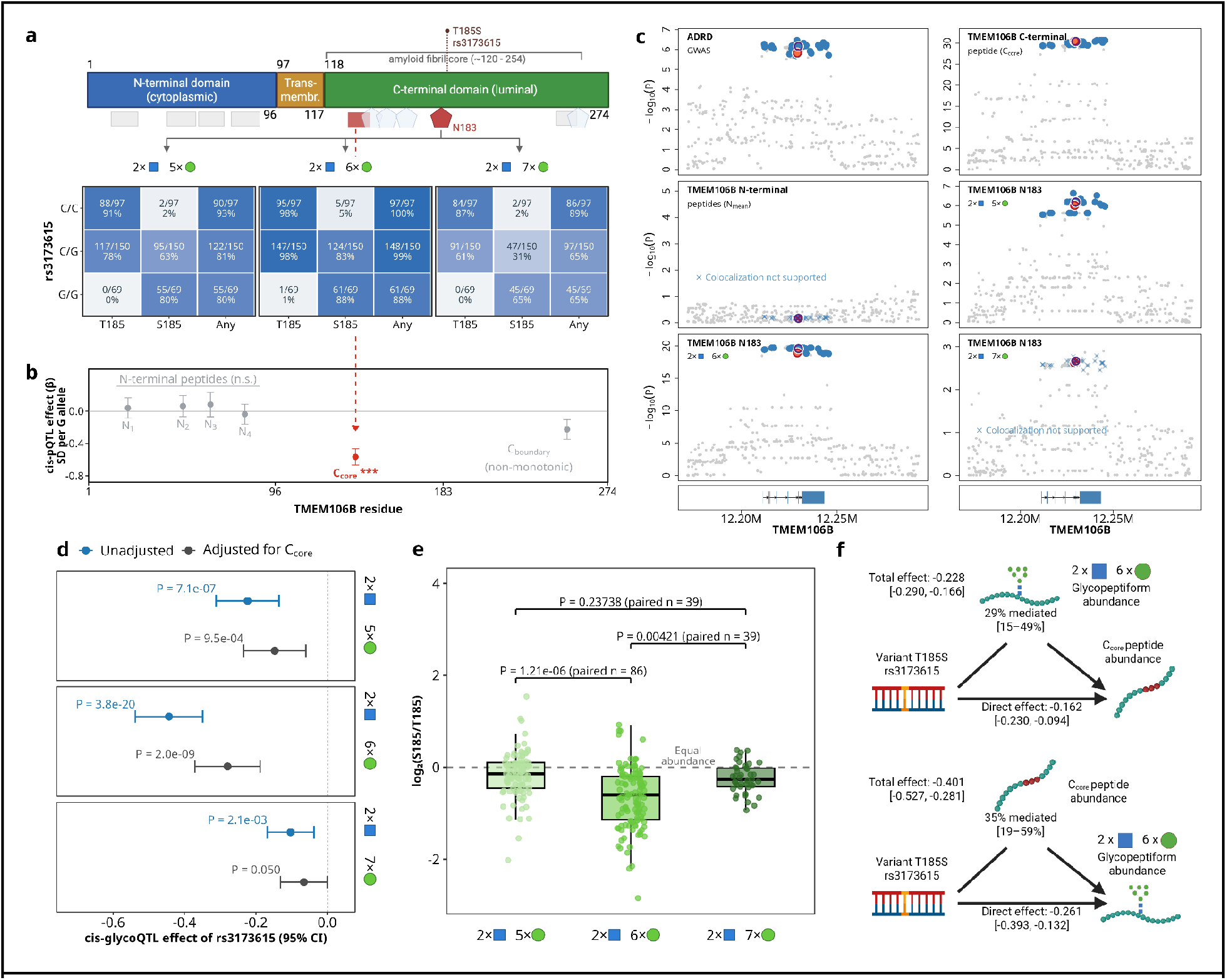
TMEM106B risk variant increases C-terminal peptide abundance and preferentially enriches HexNAc(2)Hex(6) glycopeptiforms at N183. a) Schematic of TMEM106B showing the cytoplasmic N-terminal domain, transmembrane domain, luminal C-terminal domain, five predicted N-glycosylation sites (hexagons; N183 in red), and six measured tryptic peptides (rectangles; C_core_ in red). The heatmap shows the fraction of donors with quantified T185- or S185-containing glycopeptiforms carrying HexNAc(2)Hex(5-7) at N183, stratified by rs3173615 genotype. C encodes the risk T185 allele; G encodes the protective S185 allele. b) Cis-pQTL effect of rs3173615 on each tryptic peptide (β in SD per S185/G allele, 95% CI), positioned along the protein’s amino acid coordinates. The C_core_ peptide (red arrow), located in the C-terminal domain, showed a negative effect. c) Association plots for the ADRD GWAS signal, C_core_ abundance, N-terminal peptide score (N_mean_), and the three allele-summed N183 glycopeptiform features. Each dot represents a genetic variant, with the strength of association shown as log-transformed P value. Colocalization with ADRD risk was supported for the C_core_ peptide and the allele-summed HexNAc(2)Hex(5) and HexNAc(2)Hex(6) features. Variants in the 95% colocalization confidence set are shown in blue. Coral and outlined markers indicate rs5011432 and rs3173615, respectively, which are in near-complete LD (r² = 0.988). d) Cis-glycoQTL effects of rs3173615 per S185/G allele on the three allele-summed glycopeptiform features, before adjustment for C_core_ abundance (blue) and using C_core-adjusted abundances (gray). Points indicate effect estimates and error bars indicate 95% confidence intervals. e) Within-donor log₂(S185/T185) intensity ratios for HexNAc(2)Hex(5–7) in rs3173615 C/G heterozygotes. Points represent donors with both corresponding allelic glycopeptiforms quantified. The dashed line indicates equal S185 and T185 intensity. Brackets show unadjusted P values from two-sided paired t-tests comparing log₂ ratios between glycan compositions, using donors with both ratios available. f) Mediation analyses of rs3173615, with allele-summed HexNAc(2)Hex(6) abundance and C_core_ abundance alternately specified as mediator and outcome. Total effects, indirect effects (ACME), and direct effects (ADE) are shown per S185/G allele with 95% confidence intervals.

To quantify TMEM106B glycopeptiforms across donors, we searched the mass spectra against a database containing both T185 and S185 peptide sequences. We quantified 16 TMEM106B glycopeptiforms representing six glycan compositions, all at N183 **(Table S3)**. After excluding the two compositions quantified in fewer than five donors and glycopeptiforms containing oxidized methionine, we retained four pairs of T185- and S185-containing glycopeptiforms carrying HexNAc(2)Hex(5-8) **(Fig. S4a)**. We subsequently excluded the HexNAc(2)Hex(8) pair from QTL analyses because the S185-containing glycopeptiform was poorly quantified, particularly in carriers of the protective G allele **(Fig. S4a,b)**. Quantification rates for the remaining three pairs, stratified by genotype, are shown in **Fig. 4a**. For each glycan composition, we summed the intensities of the corresponding T185 and S185 glycopeptiforms. Two of these allele-summed features, corresponding to HexNAc(2)Hex(5) and HexNAc(2)Hex(6), reached genome-wide significance in the GlycoWAS analysis, which integrated glycoQTLs with ADRD GWAS signals **(Fig. 3a; Table S2)**.

The glycosite N183 is located within the luminal C-terminal region of TMEM106B, where proteolytic cleavage generates fragments that can form amyloid fibrils in the aging human brain^49–52^. We therefore examined the previously reported rs3173615 pQTL^47^ at the peptide level, distinguishing peptides from different regions of TMEM106B. Using our TMT proteomic data, we quantified six TMEM106B peptides: four mapping to the cytoplasmic N-terminal domain (N_1_-N_4_), one within the amyloid fibril core (C_core_), and one spanning the C-terminal boundary of the structurally resolved fibril core (C_boundary_)^49^. The protective G allele was associated with lower C_core_ abundance, whereas none of the four N-terminal peptides showed a significant association **(Fig. 4b; Table S4)**. C_boundary_ also showed a significant association, although its abundance followed a non-monotonic pattern across genotypes **(Fig. S4c)**. These findings suggest that the previously reported TMEM106B pQTL primarily reflects variation in C-terminal peptide abundance, within the domain containing the N183 glycosite. For subsequent fine-mapping and colocalization of pQTLs, glycoQTLs and ADRD GWAS signals, we therefore analyzed two peptide-based abundance measures separately: the mean abundance (N_mean_) of the two most frequently detected N-terminal peptides (N_1_ and N_4_) and C_core_ abundance.

To further study the genetic mechanisms at this locus, we performed multi-context colocalization and fine-mapping^43^ of the three allele-summed glycopeptiform features, the peptide-based measures N_mean_ and C_core_, and ADRD GWAS summary statistics **(Fig. 4c)**. The multi-context analysis supports a shared genetic signal underlying ADRD risk, C_core_ abundance, and the allele-summed abundances of the HexNAc(2)Hex(5) and HexNAc(2)Hex(6) glycopeptiforms **(Table 1)**. HexNAc(2)Hex(7) showed a similar regional association pattern, but the weaker QTL evidence was insufficient to support its inclusion in the shared colocalization signal in our model. Consistent with the preceding peptide-level analysis, no significant QTL was detected for N_mean_. The 95% colocalization confidence set (CoS) contained 49 variants, reflecting the complex linkage disequilibrium (LD) structure at this locus. The lead variant was rs5011432. However, the coding variant rs3173615 (T185S) was also included in the CoS and was in strong LD with rs5011432 (r² = 0.988). Thus, our fine-mapping retains T185S as a candidate causal variant among other highly correlated variants.

**Table 1:** Genetic effects of rs3173615 (T185S) on TMEM106B glycopeptiforms.

| **Glycan composition** | **Site** | **Effect size<sup>§</sup>** | ***P* value<sup>§</sup>** | **S185/T185** |
|---|---|---|---|---|
| HexNAc(2)Hex(5)* | N183 | -0.270 [-0.370, -0.165] | 6.53 × 10<sup>−7</sup> | 0.90 [0.83, 0.97] |
| HexNAc(2)Hex(6)* | N183 | -0.462 [-0.545, -0.371] | 2.58 × 10<sup>−20</sup> | 0.63 [0.58, 0.69] |
| HexNAc(2)Hex(7) | N183 | -0.171 [-0.276, -0.061] | 0.00216 | 0.85 [0.79, 0.91] |

\* Colocalized with TMEM106B C-terminal peptide C<sub>core</sub> and ADRD GWAS signal.
<sup>§</sup> P value and effect size were estimated by ColocBoost for allele-summed glycopeptiform features.

To assess whether the glycoQTL associations persisted after accounting for C-terminal peptide abundance, we repeated the rs3173615 association analysis using C_core_-adjusted abundances of the allele-summed glycopeptiform features **(Fig. 4d)**. The associations with HexNAc(2)Hex(5) and HexNAc(2)Hex(6) remained significant, suggesting that these glycoQTL signals were not fully explained by variation in measured C_core_ abundance.

The differences in glycoQTL effect sizes **(Fig. 4d)** prompted us to test whether the imbalance between S185- and T185-containing glycopeptiforms depended on glycan composition. Among the 150 heterozygous donors, we calculated within-donor S185/T185 abundance ratios for each glycan whenever both allelic glycopeptiforms were quantified **(Fig. 4e)**. S185-containing glycopeptiforms had lower measured abundance than their T185 counterparts for all three glycan compositions, with the largest deficit observed for HexNAc(2)Hex(6). For this glycan, S185 abundance was 36.6% lower than T185 abundance (mean log₂ S185/T185 = −0.659; 95% CI, −0.774 to −0.543; n = 123). Paired comparisons among heterozygous donors with ratios available for both glycan compositions showed that the S185/T185 ratio was significantly lower for HexNAc(2)Hex(6) than for either HexNAc(2)Hex(5) or HexNAc(2)Hex(7) (P = 1.21 ×10^−6^ and 0.00421; n = 86 and 39, respectively). These within-donor comparisons reveal a glycan-dependent allelic imbalance at N183, with preferential depletion of the S185-containing HexNAc(2)Hex(6) glycopeptiform.

Finally, we performed mediation analyses of the relationship between rs3173615, HexNAc(2)Hex(6) abundance, and C_core_ abundance **(Fig. 4f)**. Significant indirect effects were estimated when HexNAc(2)Hex(6) abundance was specified as the mediator and C_core_ abundance as the outcome, but also when these roles were reversed, leaving the causal direction unresolved. Together, the genetic and within-donor analyses connect ADRD risk to increased C-terminal TMEM106B peptide and glycopeptide abundance and reveal a glycan-dependent allelic imbalance at N183, suggesting HexNAc(2)Hex(6)-carrying TMEM106B as a candidate mechanistic link between genetic risk and C-terminal fragment accumulation.

### ACE risk variant is associated with contrasting changes in glycopeptiform abundance

In addition to TMEM106B, angiotensin-converting enzyme (ACE) emerged as a significant locus in our GlycoWAS. ACE is a highly glycosylated membrane-bound zinc metallopeptidase that has previously been implicated in ADRD through vascular and β-amyloid-related mechanisms. We detected nine ACE glycopeptiforms, of which one was excluded because it was quantified in only 38% of samples, leaving eight glycopeptiforms spanning four glycosites **(Fig. 5a**, **Table 2)**. Joint multi-omics colocalization analysis including ADRD GWAS, proteomic, and glycoproteomic data supported a model in which a shared causal variant underlies the ADRD association, an ACE pQTL, and glycoQTLs of six ACE glycopeptiforms **(Fig. 5b, Fig. S5a)**. The lead variant in the 95% CoS was rs8077276.

**Figure 5.**
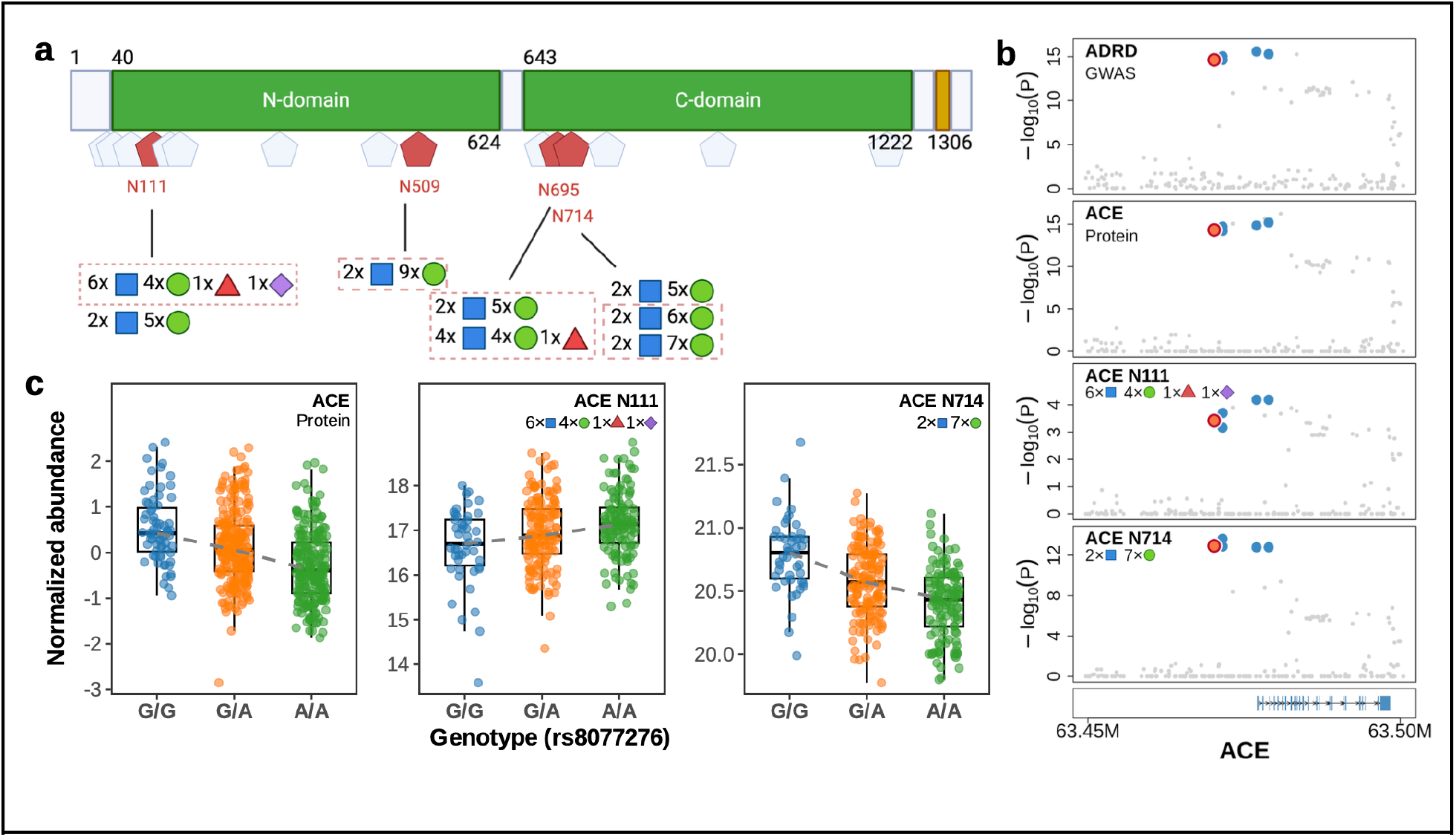
ACE risk variant is associated with contrasting changes in glycopeptiform abundance. a) Schematic of ACE showing the N-terminal and C-terminal catalytic domains, the transmembrane domain, and predicted N-glycosylation sites (hexagons). The eight detected glycopeptiforms originate from four glycosylation sites (red hexagons), with the corresponding glycan compositions shown below each site. Red dashed boxes indicate glycopeptiforms whose cis-glycoQTLs colocalized with both the ACE pQTL and AD GWAS signal. b) Association plots for the ADRD GWAS signal, ACE protein abundance (pQTL), and two ACE glycopeptiforms (HexNAc(6)Hex(4)Fuc(1)NeuAc(1) at N111 and HexNAc(2)Hex(7) at N714) at the ACE locus. Each dot represents a genetic variant, with the strength of association shown as the log-transformed P value. The ADRD GWAS signal colocalizes with the ACE pQTL and both representative glycopeptiforms. Variants in the 95% colocalization confidence set are shown in blue. The lead variant (rs8077276) is highlighted in coral with a red outline. c) ACE protein abundance and two ACE glycopeptiforms carrying HexNAc(6)Hex(4)Fuc(1)NeuAc(1) at N111 and HexNAc(2)Hex(7) at N714 stratified by rs8077276 genotype. ACE protein abundance and the N714 glycopeptiform decreased with risk allele A dosage, whereas the N111 glycopeptiform increased. Boxplots show the median and interquartile range. The dashed line connects genotype-group medians.

**Table 2:** Genetic effects of rs8077276 on ACE glycopeptiforms.

| **Glycan composition** | **Site** | **Glycan class** | **Effect size<sup>§</sup>** | ***P* value<sup>§</sup>** |
|---|---|---|---|---|
| HexNAc(2)Hex(5) | N111 | High-mannose | -0.220 [-0.323, -0.113] | 6.21 × 10<sup>−5</sup> |
| HexNAc(6)Hex(4)Fuc(1)NeuAc(1)* | N111 | Hybrid/complex | 0.197 [0.088, 0.301] | 3.78 × 10<sup>−4</sup> |
| HexNAc(2)Hex(5) | N714 | High-mannose | -0.146 [-0.253, -0.037] | 0.00871 |
| HexNAc(2)Hex(6)* | N714 | High-mannose | -0.298 [-0.395, -0.194] | 3.30 × 10<sup>−8</sup> |
| HexNAc(2)Hex(7)* | N714 | High-mannose | -0.386 [-0.476, -0.288] | 1.21 × 10<sup>−13</sup> |
| HexNAc(2)Hex(9)* | N509 | High-mannose | -0.362 [-0.454, -0.262] | 6.19 × 10<sup>−12</sup> |
| HexNAc(2)Hex(5)* | N695 | High-mannose | -0.259 [-0.359, -0.153] | 2.11 × 10<sup>−6</sup> |
| HexNAc(4)Hex(4)Fuc(1)* | N695 | Hybrid/complex | -0.326 [-0.421, -0.223] | 1.06 × 10<sup>−9</sup> |

\*Colocalized with ACE pQTL and AD GWAS signal.
<sup>§</sup> P value and effect size were estimated by ColocBoost at rs8077276.

The ADRD risk allele A was associated with lower ACE protein abundance **(Fig. 5c)**. Consistent with this observation, five of the six colocalized glycopeptiforms showed reduced abundance in risk allele carriers **(Fig. 5c, Fig. S5b)**. The two glycopeptiforms containing N111 showed comparatively modest statistical evidence of association and contrasting effect directions **(Table 2)**. The HexNAc(2)Hex(5) glycopeptiform showed lower abundance in risk allele carriers but did not meet the criteria for colocalization. In contrast, the colocalized glycopeptiform assigned the total glycan composition HexNAc(6)Hex(4)Fuc(1)NeuAc(1) showed higher abundance in risk allele carriers. For the latter, we could not distinguish a single N-linked glycan at N111 from an alternative assignment comprising both an N-linked glycan at N111 and an O-linked glycan at T113. N111 is located within the N-terminal catalytic domain of ACE, which has been implicated in β-amyloid degradation^53,54^. The biological significance of the selective increase of this glycopeptiform in risk allele carriers cannot be resolved from our data.

### Targeted trans-glycoQTL mapping identifies long-range genetic regulation of the brain glycoproteome

To explore long-range genetic influences on protein glycosylation, we performed a targeted trans-glycoQTL analysis restricted to variants previously implicated in gene regulation by external genetic studies and databases (methods). We defined trans-glycoQTLs as associations in which the genetic variant and the gene encoding the target glycopeptiform were located on different chromosomes. Using a marginal screening threshold (MAF 0.05, P < ≥) followed by fine-mapping, this analysis nominated 1,183 credible sets involving 1,088 5×10^−8^glycopeptiforms from 435 glycoproteins (Fig. 6a).

**Figure 6.**
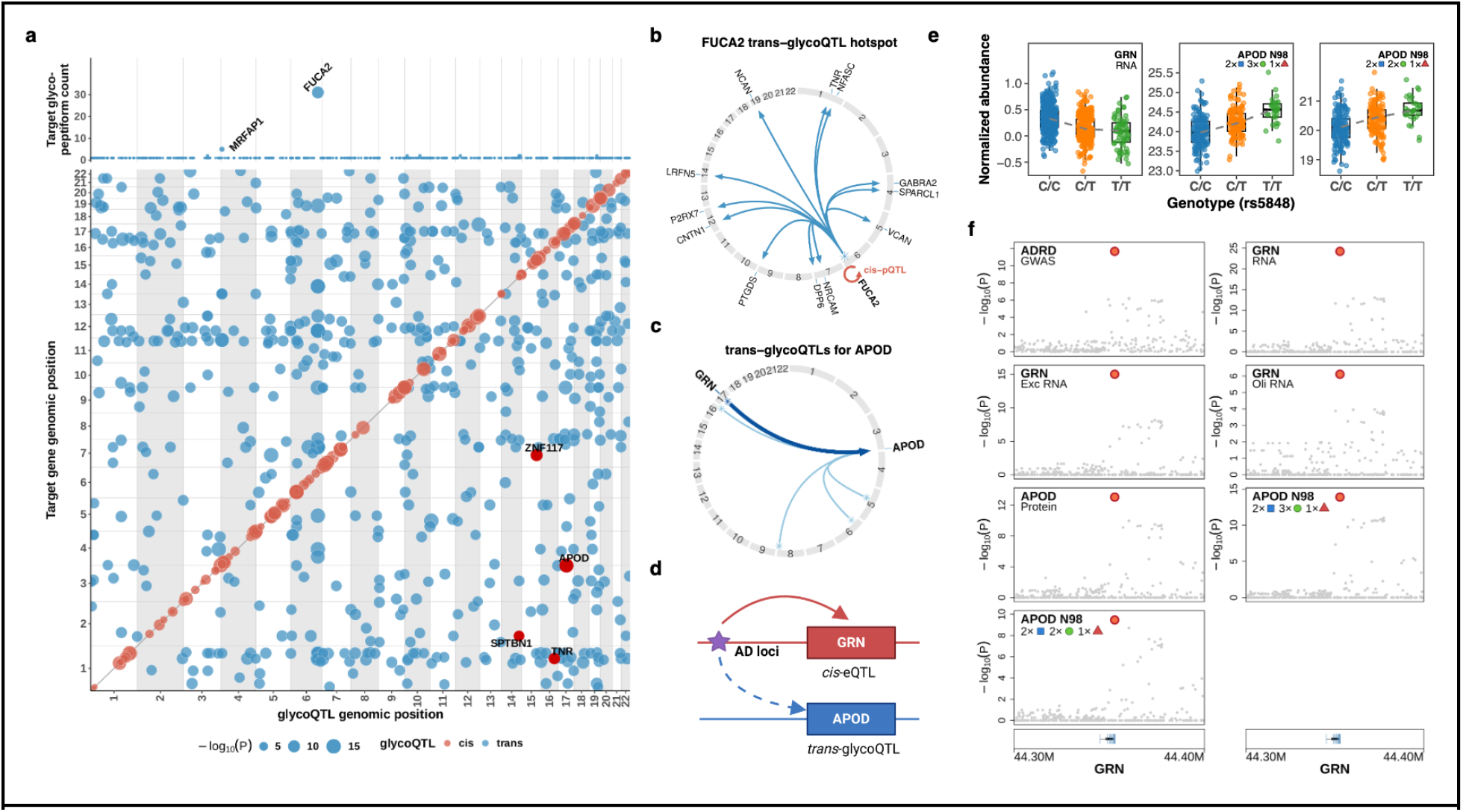
Trans genetic regulation of glycoproteins. a) Genome-wide distribution of trans-glycoQTL associations. Each blue dot represents a lead variant (x-axis) and one associated glycopeptiform, positioned by the genomic location of its corresponding gene on a different chromosome (y-axis). Dot size indicates the strength of the association. cis-glycoQTLs are shown as coral dots along the diagonal. Red dots indicate target glycoproteins encoded by known ADRD risk genes. Display filters (95% credible set size ≤ 5 and lead-variant PIP > 0.1) were applied to trans associations; all cis associations with MAF ≥ 0.05 are shown. Dots in the top panel indicate the number of glycopeptiforms associated with each trans-glycoQTL. b) Circos plot showing the FUCA2 trans-glycoQTL hotspot, represented by missense lead variant rs11155297. Blue arrows point from the glycoQTL locus to its trans target genes. The red arrow indicates the cis-pQTL association with FUCA2. c) Circos plot showing trans-glycoQTLs associated with APOD. Blue arrows point from trans-glycoQTL loci on different chromosomes to APOD. The bold blue arrow indicates the APOD trans-glycoQTL at the GRN locus. d) Schematic illustrating a model of a single shared causal variant at the GRN locus, which is associated with GRN RNA levels and APOD glycopeptiforms. e) GRN RNA abundance and APOD glycopeptiforms carrying HexNAc(2)Hex(3)Fuc(1) and HexNAc(2)Hex(2)Fuc(1) stratified by rs5848 genotype. GRN RNA abundance decreased and the two APOD paucimannose glycopeptiforms increased with risk T allele dosage. Boxplots show the median and interquartile range. The dashed line connects genotype-group medians. f) Association plots for the ADRD GWAS signal, GRN RNA abundance (bulk tissue, excitatory neurons, and oligodendrocytes), APOD protein abundance (pQTL), and the APOD glycopeptiforms carrying HexNAc(2)Hex(3)Fuc(1) and HexNAc(2)Hex(2)Fuc(1). Each dot represents a genetic variant, with the strength of association shown as the log-transformed P value. The ADRD GWAS signal colocalizes with the GRN cis-eQTLs, the APOD pQTL, and the two APOD glycoQTLs. The lead variant (rs5848), which is the only variant in the 95% colocalization confidence set, is highlighted in orange with a red outline.

We next investigated the trans-glycoQTL hotspot with the largest number of target glycopeptiforms. This locus, tagged by the missense variant rs11155297 in FUCA2, was associated with 31 glycopeptiforms from 12 glycoproteins (Fig. 6b). FUCA2 encodes ɑ-L-fucosidase 2, a member of glycoside hydrolase family 29 for which fucosidase activity has been reported, although its physiological substrates and contribution to glycan defucosylation remain uncertain55. The variant rs11155297 has previously been reported as a FUCA2 pQTL in plasma56. Consistently, our brain proteomic data also identified rs11155297 as a cis-pQTL for FUCA2 (p = 5.8 × 10^−25^) (Fig. S6a). These findings suggest that genetic variation affecting 5.8 ×10^−25^ FUCA2 abundance or activity may exert widespread effects on the brain glycoproteome.

Finally, we examined whether genes targeted by trans-glycoQTLs included established ADRD risk genes. Among the four ADRD risk genes identified as candidate trans-glycoQTL targets, APOD showed the strongest evidence for a long-range regulatory interaction **(Fig. 6a)**. APOD glycopeptiforms were associated with five independent trans-glycoQTLs, including rs5848, an established ADRD risk variant located in the 3’ untranslated region of GRN **(Fig. 6c)**. The rs5848 variant is a strong cis-eQTL for GRN and was associated in trans with APOD protein abundance as well as six paucimannose APOD glycopeptiforms **(Fig. 6d,e, S6b)**. Multi-omics colocalization supported a model with a shared causal variant underlying the ADRD GWAS signal, GRN eQTL in bulk tissue, excitatory neurons and oligodendrocytes, the APOD trans-pQTL, and six APOD trans-glycoQTLs, with rs5848 showing the highest variant colocalization probability within the 95% colocalization confidence set **(Fig. 6f)**. In contrast, the remaining APOD glycopeptiforms showed little or no evidence of colocalization with rs5848 **(Table S5, Fig. S6c)**. The corresponding GRN cis-pQTL was comparatively weak in our brain proteomic data **(Fig. S6b)**, although rs5848 has previously been identified as a GRN cis-pQTL in plasma^57^.

Thus, these targeted analyses demonstrate that trans-glycoQTL mapping can identify broad regulators of brain glycopeptiform abundance, as shown by FUCA2, and nominate disease-associated long-range regulatory interactions, as shown by the GRN-APOD locus.

## Discussion

This study set out to investigate the effect of genetic variation on the human brain glycoproteome. We generated a novel glycoproteome dataset covering N-glycosylation (7,500 glycopeptiforms) and glycation (1,582 glycopeptiforms) in 316 human cortex samples with matched whole-genome sequencing data. In contrast to previous glycome studies^25–28^, our site-specific glycoproteomic approach enabled us to investigate genetic effects on individual glycopeptiforms and glycosylation sites. We first characterized cis-glycoQTLs, defining a glycoQTL as a genetic locus associated with the abundance of a glycopeptiform. As expected, many glycoQTLs colocalized with pQTLs, indicating that genetic effects on glycopeptiform abundance may often be mediated through changes in total protein abundance. Their enrichment in regulatory regions further supports a contribution from gene regulatory mechanisms. However, many glycoQTLs remained significant after accounting for matched protein abundance, and glycosylated peptides were more frequently regulated by genetic variants than glycated peptides, suggesting that common genetic variation influences glycosylation beyond its effects on steady-state protein abundance.

Interestingly, glycoQTLs were enriched among high-mannose compared with complex or hybrid glycopeptiforms, indicating that genetic regulation may preferentially affect early steps of N-glycan biosynthesis in the ER, including co-translational glycan attachment and initial glycan processing. One possible explanation is that non-coding regulatory variants alter the rate of protein production or trafficking through the secretory pathway, thereby changing exposure to glycosylation enzymes without necessarily producing detectable changes in steady-state protein abundance. Consistent with this mechanism, previous studies have shown that co-translational processes, including translation kinetics, can affect N-linked glycosylation efficiency^58^. Our findings suggest that this concept extends to common human genetic variation and that regulatory variants can influence glycoproteoform composition through subtle changes in protein biogenesis rather than solely through changes in steady-state protein abundance.

After establishing a glycoproteome-wide cis-QTL map of the human brain, we integrated these data with ADRD GWAS summary statistics to identify glycoQTLs that are associated with disease risk. We found five genes with colocalized glycoQTL and ADRD risk signals, among which TMEM106B emerged as perhaps the most intriguing for further investigation. TMEM106B has been intensively studied in the context of frontotemporal dementia, but the locus has also been identified in recent ADRD GWAS^3,4^. The ADRD risk variant has previously been associated with increased TMEM106B protein abundance^47^, whereas evidence for an eQTL effect is relatively weak. Recent independent studies demonstrated that a C-terminal fragment (CTF), spanning almost the entire luminal C-domain, can form amyloid-like fibrils that accumulate in the aging human brain^49–51^. The glycosite N183 detected in our dataset is located within this CTF and is of particular interest because the coding variant T185S, a candidate causal variant supported by our fine-mapping, directly alters its glycosylation sequon, converting the generally preferred N-X-T to N-X-S^48,59^. Glycosylation at N183 has been confirmed by biochemical assays^46^ and, more recently, visualized in brain-derived amyloid fibrils by cryo-EM^49^. However, the glycan compositions present at this site in the human brain and their relationship to T185S remained incompletely characterized^47^.

Our mass-spectrometry approach identified six distinct glycan-associated mass shifts at N183, of which three, corresponding to HexNAc(2)Hex(5-7), could be quantified reliably for allele-specific analysis. Across these three glycan compositions, S185-containing glycopeptiforms showed lower measured abundances than their T185 counterparts. Particularly interesting was the variation in allelic ratios across glycan compositions within heterozygous donors: the strongest relative depletion of S185 was observed for the HexNAc(2)Hex(6)-carrying glycopeptiform. Although T185- and S185-containing glycopeptides could potentially differ in ionization or detection efficiency, our observation within heterozygous donors supports a glycan-dependent association between T185S and glycopeptiform abundance at N183. Thus, our work characterized the glycan compositions at N183, demonstrated glycan-dependent differences in allelic ratios, and prioritized the HexNAc(2)Hex(6)-carrying glycopeptiform for mechanistic investigation.

Given the reported aggregation of the TMEM106B CTF, we examined genetic associations at the peptide level. The risk-associated T185 allele was associated with higher abundance of a peptide mapping to the CTF core (C_core_), whereas N-terminal peptides showed no significant association. This pattern suggests that the pQTL reflects CTF accumulation rather than a uniform change in full-length TMEM106B abundance^60^. The ADRD risk signal colocalized with both the HexNAc(2)Hex(6) glycoQTL and the C_core_ pQTL, supporting a shared genetic basis for these molecular traits and disease susceptibility. The glycoQTL remained significant after adjustment for C_core_ abundance, supporting an association with N183 glycosylation beyond variation captured by this peptide. These findings are consistent with a model in which T185-associated enrichment of the HexNAc(2)Hex(6) glycoform promotes CTF stability or aggregation. However, mediation analysis did not resolve the direction of this relationship. Alternatively, HexNAc(2)Hex(6)-carrying species could become enriched through preferential retention within the accumulating CTF pool. Our measurements also cannot distinguish glycopeptides derived from full-length TMEM106B versus cleaved CTF, or from soluble versus aggregated species. In summary, our glycoQTL study identified N183 glycan composition as a candidate molecular link between TMEM106B genetic risk and CTF accumulation, providing a target for mechanistic investigation.

In addition to TMEM106B, the ACE locus demonstrated that genetic associations with individual glycopeptiforms can diverge from those with total protein abundance. While the risk allele was associated with lower ACE protein abundance and, consistently, with lower abundances of most ACE glycopeptiforms, one colocalized glycopeptiform with a hybrid/complex glycan at N111 showed a comparatively modest QTL effect in the opposite direction. N111 is located within the N-terminal catalytic domain, which has been implicated in β-amyloid degradation^53,54^. Although the site lies away from the catalytic center, structural studies have also identified glycan-glycan contacts at this site across the ACE dimer interface^61,62^. However, the functional consequences of the observed glycosylation changes remain unknown.

Finally, we performed an exploratory trans-glycoQTL analysis to assess long-range genetic regulation of the brain glycoproteome. Since our sample size was modest for trans-QTL discovery, we adopted a targeted strategy and focused on the strongest associations. The trans-glycoQTL hotspot with the largest number of target glycopeptiforms was tagged by a missense variant in the fucosidase FUCA2, suggesting that the associated glycopeptiforms represent direct or indirect downstream targets of FUCA2. These findings provide human genetic evidence for a role of FUCA2 in glycoproteome regulation, while the underlying biochemical mechanism remains unresolved^55^. More generally, these results illustrate how larger future trans-glycoQTL studies could systematically identify the target proteins and precise glycosylation sites altered by glycan-modifying enzymes whose abundance or activity is under genetic control. Conversely, APOD emerged as the ADRD glycoprotein targeted by the largest number of independent trans-glycoQTLs. At the GRN risk locus, colocalization supported a shared genetic signal linking ADRD risk, GRN expression, APOD protein abundance, and six paucimannose APOD glycopeptiforms. This connection is biologically plausible given the involvement of both progranulin and APOD in lysosomal function^63^. Whether GRN mediates these trans-effects remains to be determined. A complementary analysis of this glycoproteomic resource identified colocalization between a QTL for an astrocyte transcriptomic program and multiple trans-glycoQTLs^64^, illustrating how this resource can help place tissue-level glycoQTLs within cell-type-resolved regulatory networks.

The broad glycopeptiform coverage provided by our mass-spectrometry approach is a major strength of this study, but the assay also has several limitations. First, despite the large number of quantified glycopeptiforms, these likely represent only a fraction of the molecular diversity of the human brain N-glycoproteome. Whether a glycopeptiform is detected depends on its abundance, enrichment efficiency, and mass-spectrometric detectability. Further, our enrichment strategy targeted only N-linked, not O-linked glycopeptides. Second, the glycoproteome was quantified at the tissue level, precluding direct assignment of glycoQTLs to specific cell types. Although glycoQTLs colocalized with eQTLs detected across all major brain cell types, the exact cellular origin of glycoQTLs remains unresolved. Differences in glycopeptiform abundance across cell types, combined with cell-type-specific genetic effects, could contribute to the divergent associations observed for different glycopeptiforms of the same protein at the tissue level. Third, our approach infers glycan composition but not glycan structure, limiting mechanistic interpretation of the functional consequences of altered glycosylation. In addition, peptide-level measurements do not reveal how glycans at different sites are combined on individual protein molecules, preventing reconstruction of intact glycoproteoforms.

Overall, we comprehensively characterized the genetic regulation of the human brain glycoproteome. By integrating site-specific glycoproteomics with proteomic, transcriptomic and genetic data, we demonstrated that common genetic variation shapes glycoproteoform composition, revealing a layer of genetic regulation not captured by protein abundance alone. These analyses further revealed molecular consequences of ADRD risk variants that would not be resolved by conventional transcriptomic or protein-level analyses, as illustrated by TMEM106B and ACE. As large-scale glycoproteomic datasets become available across tissues, cell types, and diseases, glycoQTL mapping should provide an additional molecular layer for understanding the functional consequences of human genetic variation.

## Online Methods

### ROSMAP cohort and neuropathologic characterization

Dorsolateral prefrontal cortex (DLPFC) tissue was obtained from 316 decedents of the Religious Orders Study (ROS) and Rush Memory and Aging Project (MAP), two longitudinal cohort studies of aging and dementia with harmonized clinical and postmortem data collection.^38^ At enrollment, participants were free of known dementia and agreed to annual clinical assessments and brain donation at death. Participant characteristics are summarized in **Table S1**. QTL analyses were adjusted for quantitative measures of amyloid and tau pathology. β-amyloid burden was defined as the mean square-root-transformed β-amyloid load across eight brain regions, quantified by immunohistochemistry using the 4G8 antibody and digital image analysis as the percentage of tissue area positive for amyloid. PHF-tau tangle density was similarly defined as the mean square-root-transformed density of PHF-tau tangles across the same eight regions, quantified by AT8 immunohistochemistry and digital image analysis as tangle density (counts per mm²)^65^. The ROS and MAP studies were approved by an Institutional Review Board of Rush University Medical Center. All participants signed an informed consent form, an Anatomic Gift Act for brain donation, and a repository consent to allow their data and biospecimens to be shared.

### Glycoproteomic data

Glycoproteomic profiling of dorsolateral prefrontal cortex (DLPFC) tissue from the ROS/MAP cohort was performed using a liquid chromatography-tandem mass spectrometry (LC-MS/MS)-based workflow. Briefly, tissue lysates were prepared by bead-beating, proteins were digested using the S-Trap method, and glycopeptides were enriched as described previously^39^. Glycopeptide mixtures were analyzed by nano-liquid chromatography (nanoAcquity, Waters) coupled to high-resolution tandem mass spectrometry (Orbitrap HF-X, Thermo Fisher Scientific) operated in data-dependent acquisition mode with stepped higher-energy collisional dissociation (HCD) fragmentation. Raw data were searched against the human proteome and a glycan library using Byonic^66^. Glycopeptide identifications were filtered at a 1% false discovery rate (FDR), followed by an additional minimum Byonic score threshold of 300 for increased stringency^67^. Glycopeptides were quantified using FlashLFQ with normalization and match-between-runs enabled and were subsequently log-transformed^68^. Glycopeptiforms with valid measurements in fewer than two-thirds of samples were excluded. Outlier samples with numbers of missing measurements above the 95th percentile of the missing-value distribution (3,275 missing values) were also excluded, removing 17 samples. The resulting dataset comprised 9,100 glycopeptiforms across 361 samples, including 316 samples with matched WGS and proteomic data. Manual review of noncanonical modification assignments identified two glycopeptiforms with modifications assigned to serine or threonine and 54 glycopeptiforms with putative W-linked glycans. Since these glycopeptiforms could not be confidently classified, the results reported here were restricted to the 9,044 glycopeptiforms carrying N-linked glycans or glycated lysines. For the QTL analyses, missing values were imputed using empirical Bayes matrix factorization implemented in flashier with point-Laplace priors and iterative backfitting for up to 300 iterations. The model was initialized using PCA components derived from a softImpute-completed matrix (rank = 50, λ = 30)^69^.

For QTL analyses, we applied an additional filter to exclude potential artifacts caused by coding variants that alter peptide mass. Glycopeptiforms were excluded when (i) a coding variant located within the measured peptide was present in the 95% credible set and (ii) non-imputed glycopeptiform abundances followed the genotype-dependent pattern AA > AB > BB in the raw, unadjusted data or fewer than three BB donors were available to assess this pattern. Here, A denotes the allele matching the searched peptide sequence and B the alternative coding allele. This filter removed 21 glycopeptiforms. For TMEM106B, which was affected by this filter, we repeated the database search with both the T185 and S185 peptide sequences included. For each donor and glycan composition, we summed the normalized linear intensities of the corresponding T185- and S185-containing glycopeptiforms and then applied a log₂ transformation. If only one allelic glycopeptiform was quantified (for example, in homozygous donors), its intensity was retained. If neither was quantified, the combined feature was treated as missing. The resulting allele-summed TMEM106B features were included in the QTL analyses.

N-linked glycans were classified into four classes based on glycan composition: paucimannose (truncated mannose-type glycans; HexNAc(2)Hex(1-4), with or without fucosylation), high-mannose (oligomannose-type glycans; HexNAc(2)Hex(≥5)), hybrid/complex (HexNAc(≥3)Hex(≥3), potentially with additional fucosylation and/or sialylation), and other (compositions not fitting these categories, including small or partially processed glycans). Hybrid and complex glycans were combined because their structures could not be distinguished based on glycan composition alone.

### Proteomic data

Proteomic profiling of DLPFC tissue from the ROSMAP cohort was previously performed using tandem mass tag (TMT) isobaric labeling followed by liquid chromatography-tandem mass spectrometry and has been described elsewhere^70,71^. Peptide spectral matches were filtered at <1% FDR using Percolator implemented in Thermo Proteome Discoverer^72^. Peptides were grouped using parsimony, unique peptides were used for protein-level quantification, batch effects were mitigated using pooled reference channels and median polish normalization, and protein abundances were log_2_-transformed and median-centered. Proteins with more than 40% missing values were excluded, resulting in 7,389 proteins quantified across 596 ROSMAP participants, including 316 participants with matched glycoproteomic and WGS data. Missing values were imputed using empirical Bayes matrix factorization implemented in flashier with point-Laplace priors and iterative backfitting for up to 300 iterations, initialized using PCA components derived from a softImpute-completed matrix (rank = 50, λ = 30)^69^. The resulting imputed protein abundance matrix was used for downstream analyses.

### Adjustment for protein abundance

To adjust glycopeptiform abundances for total protein abundance, glycopeptiforms were first mapped to proteins. For glycopeptiforms mapping to multiple proteins, the protein exhibiting the highest Pearson correlation with glycopeptiform abundance was selected. Of the 9,100 quantified glycopeptiforms, 134 mapped to multiple proteins, for which the candidate protein with the highest Pearson correlation was selected. Glycopeptiforms whose corresponding protein was not confidently quantified in the proteomic dataset (n=372) were excluded from the adjusted analyses. Protein adjustment was performed in the 354 samples with matched glycoproteomic and TMT proteomic data. For each glycopeptiform, glycopeptiform abundance was regressed on the abundance of the corresponding protein, and the fitted intercept plus the residuals were retained as the protein-adjusted abundance. The regression used imputed proteomic data and non-imputed glycoproteomic data. The resulting protein-adjusted matrix was subsequently imputed using the same low-rank matrix completion framework described above, based on softImpute (rank=50, λ=30) followed by empirical Bayes matrix factorization (flasiher) with point-Laplace priors and iterative backfitting (up to 300 iterations)^69^. A total of 316 participants with matched WGS data were included in the protein-adjusted analyses. Unless otherwise stated, all primary analyses used the unadjusted glycopeptiform abundances, whereas protein-adjusted abundances were used only for secondary analyses.

### RNA data processing

Single-nucleus RNA sequencing (snRNA-seq) data from DLPFC tissue of 479 ROSMAP participants were generated and processed as previously described^10,73^. Briefly, nuclei were profiled using the 10x Genomics Chromium platform, followed by doublet detection, cell-type annotation, and aggregation into donor-level pseudobulk expression profiles for six major brain cell types (excitatory neurons, inhibitory neurons, astrocytes, microglia, oligodendrocytes, and oligodendrocyte precursor cells). Expression values were normalized as counts per million (CPM) and subsequently quantile normalized.

Bulk RNA-seq data from postmortem DLPFC tissue of 1,093 ROSMAP participants were generated as previously described^74,75,75^. For this study, gene-level expression was quantified with RNA-SeQC. Sample-level quality control followed the GTEx v8 protocol, removing outliers based on relative log expression, the D-statistic summarizing average correlation between sample pairs, and the Mahalanobis distance of each sample to a hierarchical clustering of samples. Genes with TPM below 0.1 in more than 20% of samples were excluded, and genes were further required to have at least 6 reads in at least 20% of samples. Counts were then normalized with TMM.

### Genotype and Covariate processing

Whole-genome sequencing (WGS) libraries were prepared and sequenced as described previously^74^. Sequence reads were aligned to the GRCh38 reference genome, and variants were detected using the GATK pipeline^75,76^. Project VCF files were processed with bcftools: multi-allelic sites were split and variants annotated with dbSNP, then filtered by FILTER status, genotype depth (DP ≥ 10), genotype quality (GQ ≥ 20) and allele balance, and monomorphic sites were removed. Variants were then converted to PLINK^77^, followed by an additional round of QC. Samples with >10% missing genotypes were excluded, and variants deviating from Hardy-Weinberg equilibrium (*P* < 1 × 10^−8^) were removed. No minor allele frequency filter was applied. The QC-passed BED file served as the genotype input for downstream analyses.

For each molecular phenotype, we used a common covariate set consisting of sex, age at death, study (ROS or MAP), the top 15 genotype principal components, estimated hidden factors, and two measures of AD pathology: β-amyloid load and PHF-tau tangle density. The hidden factors were estimated from the processed phenotype matrix by principal component analysis (PCA)^78^, with the number of retained components determined using the Marchenko-Pastur distribution.

### Fine-mapping analysis

We performed genome-wide fine-mapping across unadjusted and protein-adjusted glycoproteomic and proteomic traits using SuSiE^40^, supplying covariate-residualized individual-level phenotype and genotype data. For each gene, we defined the analysis region as the union of the topologically associating domain (TAD) containing the gene^79^ and a window extending approximately 1 Mb beyond the gene’s transcription start and end sites, consistent with the FunGen-xQTL consortium fine-mapping framework^80^. We restricted fine-mapping to variants with a minor allele count (MAC) ≥ 5. For all molecular phenotypes, we applied an adaptive pre-screening step before running full fine-mapping using a single-effect SuSiE model (L = 1). Regions with insufficient evidence of genetic association were excluded from subsequent fine-mapping. This procedure yielded posterior inclusion probabilities (PIP) and 95% credible sets (CS) for each locus, and additionally generated cis-xQTL prediction weights for downstream TWAS analyses.

Fine-mapping results were post-processed to filter low-confidence credible sets. Specifically, we retained only credible sets meeting a minimum absolute coverage threshold of 0.8 and containing at least one variant with a posterior inclusion probability (PIP) > 0.025. Variants with a MAF < 0.05 were then removed from the resulting glycoQTL and pQTL credible sets.

### TensorQTL association analysis

We performed genome-wide cis-xQTL association testing using TensorQTL^41^. For each molecular trait, we tested all variants within the TAD-based cis region defined above, restricted to those with a MAF ≥ 0.05, using the same individual-level genotypes as in the fine-mapping analysis. Nominal association statistics were obtained from a linear model of the molecular trait on variant dosage, with the covariate set described above included as fixed effects.

To assess trait-level significance while accounting for multiple testing across variants, we permuted phenotype values across individuals to generate a null distribution of the minimum nominal p-value within the cis window, and approximated this null distribution by a beta distribution to obtain a per-trait permutation P value. We then controlled the false discovery rate across traits and considered regional cis-associations significant at q < 0.05.

### Glycoproteome- and proteome-wide association studies

We performed glycoproteome-wide and proteome-wide association studies (GlycoWAS and PWAS) to evaluate associations with ADRD risk, using the transcriptome-wide association study (TWAS) framework. Before model training, we restricted variants to MAF ≥ 0.05. For each molecular trait, we partitioned the genome into the same regions used for fine-mapping and trained prediction models for all genes whose transcription start sites fell within a given interval. At loci containing large numbers of variants, we sampled up to 8,000 variants per cross-validation fold to reduce computational burden. We used summary statistics from a recent ADRD meta-analysis^3^, which were quality-controlled using allele harmonization and LD mismatch detection with SLALOM, excluding flagged variants.

We trained six models, including Elastic Net^81^, LASSO^82^, BayesR^83^, Bayesian LASSO^84^, Sum of Single Effects (SuSiE)^40^, and Mr.ASH^85^, to estimate cis-xQTL weights for each candidate gene. Elastic Net combines L1 and L2 penalties, whereas LASSO uses only the L1 norm. Bayesian LASSO places a double-exponential prior on variant effects, and BayesR places a mixture-of-normals prior with a point mass at zero, propagating uncertainty into the weight estimates. SuSiE models the effects of multiple causal variants using a probabilistic framework, and Mr.ASH fits a multiple regression with an adaptive shrinkage prior on variant effects. We assessed predictive performance by five-fold cross-validation, retaining genes imputable in at least one method (CV R² ≥ 0.01, CV P < 0.05) and selecting the model with the highest CV R². For each retained model, we computed a TWAS Z-score from the *cis*-xQTL weights, GWAS Z-scores, and LD matrices from the ADSP R4 European-ancestry LD reference partitioned into LD blocks defined in previous work^86^. A GlycoWAS or PWAS signal was considered significant if (1) the best prediction model yielded an association P value below the Bonferroni-corrected threshold, or (2) at least half of the prediction models yielded P values below this threshold.

### Mendelian randomization

We performed Mendelian randomization (MR) using the credible-set-aware Wald-ratio approach, using fine-mapped glycoQTL and pQTL variants as instruments. For each molecular trait, defined as a glycopeptiform in the glycoproteome or a protein in the proteome, we matched instruments to the ADRD GWAS summary statistics by chromosome, position, and alleles, flipping the sign of the exposure effect when reference and alternate alleles were swapped, and derived GWAS effect sizes and standard errors from Z-scores, allele frequencies, and sample sizes under a standardized-phenotype model. We rescaled exposure effects to unit standard error. We grouped instruments by credible set and retained credible sets with cumulative PIP (cPIP) ≥ 0.5. Within each retained set, we computed a composite Wald ratio as the PIP-weighted mean of the per-variant ratios, with a delta-method standard error derived from the corresponding PIP-weighted second moment. We then obtained gene-level effects by fixed-effect inverse-variance-weighted meta-analysis across credible sets, assessing between-credible-set heterogeneity with Cochran’s Q and I². Traits with no credible set meeting the cPIP threshold were reported as non-estimable. MR associations were considered significant when a molecular trait passed Bonferroni correction for the number of molecular traits tested (meta-P < 0.05 / N), showed strong causal support with a cumulative posterior inclusion probability (cPIP) of at least 0.7, contained at least one credible set, and showed no evidence of heterogeneity, defined by Cochran’s Q P > 0.01 and I² < 0.4.

### Trans-glycoQTL analysis

We compiled a target variant set from published large-scale QTL resources and from our FunGen-xQTL flagship analyses across molecular modalities^80^. We LD-pruned each resource separately (r² > 0.64, MAF > 0.01) using ROSMAP genotypes and merged the pruned sets with the ADRD variant list to form a combined panel. We tested candidate SNP-gene associations with TensorQTL^41^ on covariate-residualized genotypes and molecular phenotypes, considering associations at *P* < 5 × 10^−6^. We assigned each SNP to an LD block by genomic coordinate and annotated each block with its associated genes, yielding a block-gene map that constrained the search space for trans fine-mapping. We then fine-mapped each block with SuSiE^40^, using the LD block as the analysis window, to generate genome-wide trans-glycoQTL results. Among credible-set variants we retained those with MAF ≥ 0.05 and marginal association *P* < 5 × 10^−8^, computed from the univariate effect estimate and its standard error.

### Colocalization analysis

We performed three multi-context colocalization analyses to evaluate genetic signals shared between glycoproteomic traits and other molecular traits or ADRD risk using ColocBoost^43^. Each colocalization event was summarized by a 95% colocalization confidence set for the shared causal variants, and a corresponding trait-sharing configuration specifying which traits shared this signal. First, we performed a genome-wide colocalization analysis, which jointly analyzed glycoproteomic and expression traits to identify cell-type-level cis-eQTL signals that colocalized with tissue-level glycoQTL for each gene. Second, we jointly analyzed glycoproteomic and proteomic traits mapped to the same gene, together with ADRD GWAS summary statistics, using the GWAS-prioritized mode. This analysis identified glycoQTL that colocalized with ADRD association signals genome-wide, thereby separating signals shared with the corresponding pQTL from glycopeptiform-specific effects undetected at the protein level. Third, at two loci prioritized by trans-glycoQTL mapping, we performed targeted cis-trans colocalization to characterize shared genetic signals underlying long-range molecular associations. At each locus, we jointly analyzed the prioritized trans-associated glycopeptiforms with cis expression and proteomic traits linked to the same variant; ADRD GWAS summary statistics were also included when the trans-glycoQTL was associated with ADRD risk. All three analyses used the TAD-based cis region defined for the fine-mapping analysis. The genome-wide analysis was restricted to variants with MAF ≥0.05, while targets TMEM106B, ACE, GRN/APOD used the default MAC ≥5 threshold. Consistent with the other analyses, we used the covariate-residualized individual-level phenotype and genotype data for molecular traits, and used the quality-controlled ADRD GWAS summary statistics and LD reference panel for analyses involving ADRD. ColocBoost was run with default settings, and we retained only colocalization events containing at least one variant with marginal *P* < 1 × 10^−4^.

### Excess of overlap (EOO) analysis

We evaluated the enrichment of genomic annotations among significant fine-mapped variants (PIP > 0.1) within 95% credible sets using the excess-of-overlap framework described previously^87^. Variants in the baseline annotation panel served as the background, and significance was assessed using a leave-one-chromosome-out (LOCO) block jackknife framework. We compiled annotations from FILER^88^ and represented them as binary indicators in variant-level annotation matrices. For each annotation, we computed an odds ratio contrasting annotated and unannotated variants by significance status, and an enrichment ratio defined as the proportion of significant variants falling within the annotation divided by the proportion of all background variants in that annotation. Because variants on the same chromosome are correlated through linkage disequilibrium, we recomputed both statistics 22 times, excluding one autosome at a time, and reported the mean log₂ odds ratio and mean enrichment ratio across replicates with jackknife standard errors derived from their between-replicate variability. We compared enrichment profiles across datasets using a two-sample z-test on the difference in enrichment, restricted to annotations with P < 0.05.

### Heritability enrichment analysis

We used stratified linkage disequilibrium score regression (S-LDSC) implemented in the polyfun^89^ package to quantify the contribution of our annotations to complex trait heritability, following the procedure described previously^87^. We encoded a binary annotation recording whether a SNP belonged to our significant glycosylation glycoQTL variant set, defined as fine-mapped variants with PIP > 0.1 and MAF ≥ 0.05 within 95% credible sets. The annotation was analyzed conditional on the 97-category baseline-LD v2.2 model, using LD scores computed from the ADSP reference panel and publicly available GWAS summary statistics for 27 traits^1–4,44,90–96^. We report the standardized effect size τ*, the proportionate change in per-SNP heritability per one-standard-deviation increase in the annotation, with standard errors from the 200-block LDSC jackknife.

### Mediation analysis

To test whether the genetic effect on TMEM106B protein abundance acts through glycosylation, we performed causal mediation analysis with the R package mediation^97^. For the target locus, we used the dosage of the Ser185 (G) allele as the exposure. In the forward model, the mediator was the abundance of the HexNAc(2)Hex(6) glycopeptiform and the outcome was the abundance of the C_core_ peptide. In the switched model, C_core_ was the mediator and the HexNAc(2)Hex(6) glycopeptiform was the outcome. For each direction, we fit a mediator model (mediator ∼ exposure + covariates) and an outcome model (outcome ∼ mediator + exposure + covariates), both adjusting for β-amyloid load, PHF-tau tangle density, neocortical TDP-43 pathology (presence or absence of cortical TDP-43 pathology determined by neocortical phosphorylated monoclonal TAR5P-1D3 staining), age at death, and sex. We estimated the average causal mediation effect (ACME), average direct effect (ADE), total effect, and proportion mediated, with 95% confidence intervals and P values from 5,000 nonparametric bootstrap replicates. Effects are expressed per Ser185 allele.

### Data and code availability

The glycoproteomic data are available via the PRIDE repository under accession PXD072371. QTL and colocalization results are available via Synapse: glycoQTLs (syn76495192), multi-omics colocalization with ADRD GWAS (syn76490205), colocalization with pQTLs and cell-type-specific eQTLs (syn76489398). The proteomic, WGS and single-nucleus RNA-seq data can be accessed via the AD Knowledge Portal (https://adknowledgeportal.org).

Pathologic and phenotypic data from the ROSMAP cohort are available upon request via our Resource Sharing Hub at https://www.radc.rush.edu.

All analysis scripts, including those used for fine-mapping, multi-omics colocalization, GlycoWAS, and generation of the manuscript figures, are available on GitHub: https://github.com/rfeng2023/glycoQTL

## Data Availability

The glycoproteomic data are available via the PRIDE repository under accession PXD072371. QTL and colocalization results are available via Synapse: glycoQTLs (syn76495192), multi-omics colocalization with ADRD GWAS (syn76490205), colocalization with pQTLs and cell-type-specific eQTLs (syn76489398). The proteomic, WGS and single-nucleus RNA-seq data can be accessed via the AD Knowledge Portal (https://adknowledgeportal.org).
Pathologic and phenotypic data from the ROSMAP cohort are available upon request via our Resource Sharing Hub at https://www.radc.rush.edu.

https://doi.org/10.6019/PXD072371

https://www.radc.rush.edu

## Acknowledgements

We thank the participants of the Religious Orders Study and Rush Memory and Aging Project (ROSMAP) for their invaluable contributions, including the donation of brain tissue that made this research possible.

## Author contributions

R.F., A.W., X.C., S.S., A.P., R.L., H.S., Y. Li, K.D. and H.K. analyzed the data. D.M., H.L. and Y. Levin generated the processed glycoproteomic data. N.S. provided the proteomic data. D.B. provided ROSMAP cohort data and resources. P.D. and G.W. provided FunGen-xQTL resources. M.B., A.B. and H.K. conceived and designed the project. G.W. and H.K. developed the analysis plan. R.F. and H.K. wrote the manuscript with input from all authors. All authors interpreted and discussed the results.

