## Supplementary Figures for "Genetic architecture of the human brain glycoproteome implicates glycosylation in Alzheimer’s disease and related dementias"

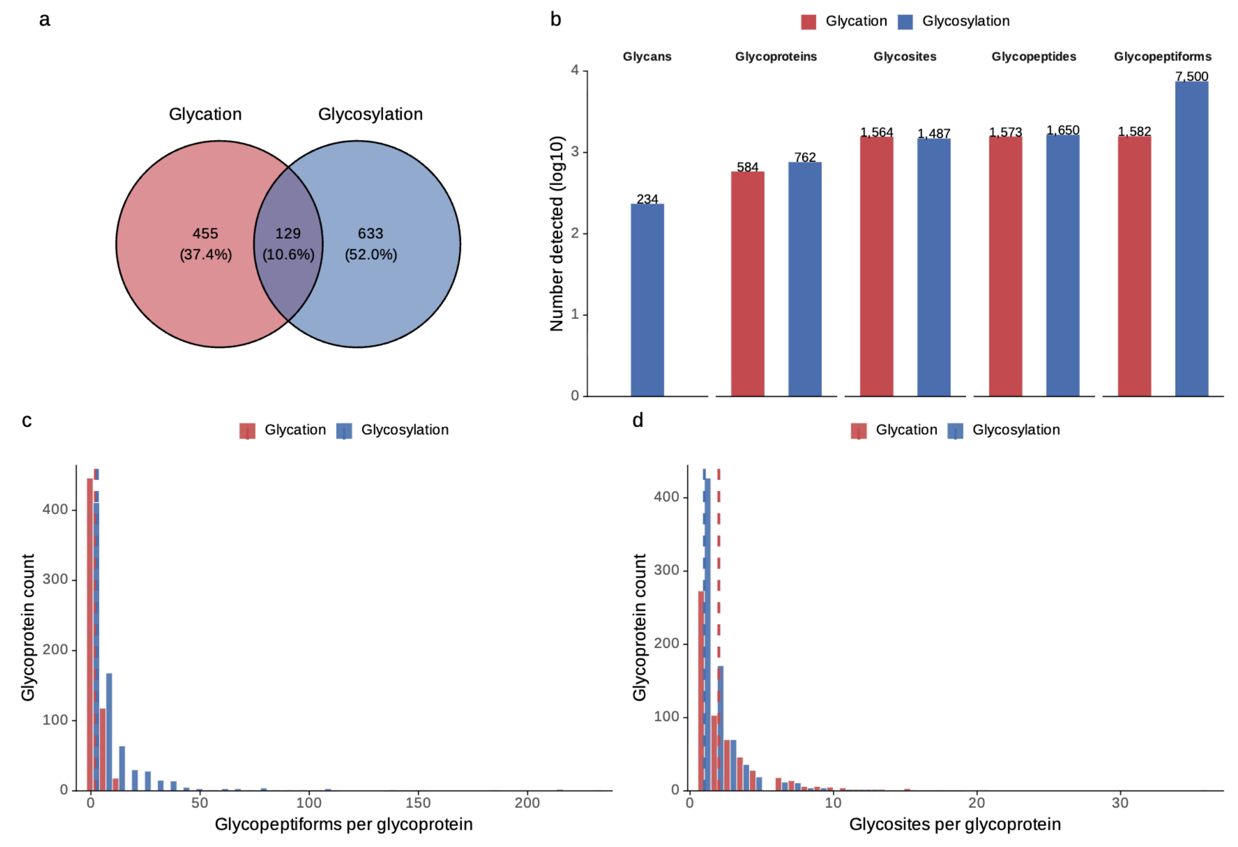


**Supplementary Figure 1. Additional characterization of the human brain glycoproteome.**
a) Venn diagram showing the numbers of detected glycated and N-glycosylated glycoproteins. The intersection represents proteins detected with both glycation and N-linked glycosylation.
b) Total number of detected glycan compositions, glycoproteins, glycosites, glycopeptides, and glycopeptiforms in the dataset.
c) Distribution of detected glycopeptiforms per glycoprotein for glycation and N-glycosylation. Vertical dashed lines indicate the median for each group.
d) Distribution of detected glycosites per glycoprotein for glycation and N-glycosylation. Vertical dashed lines indicate the median for each group.

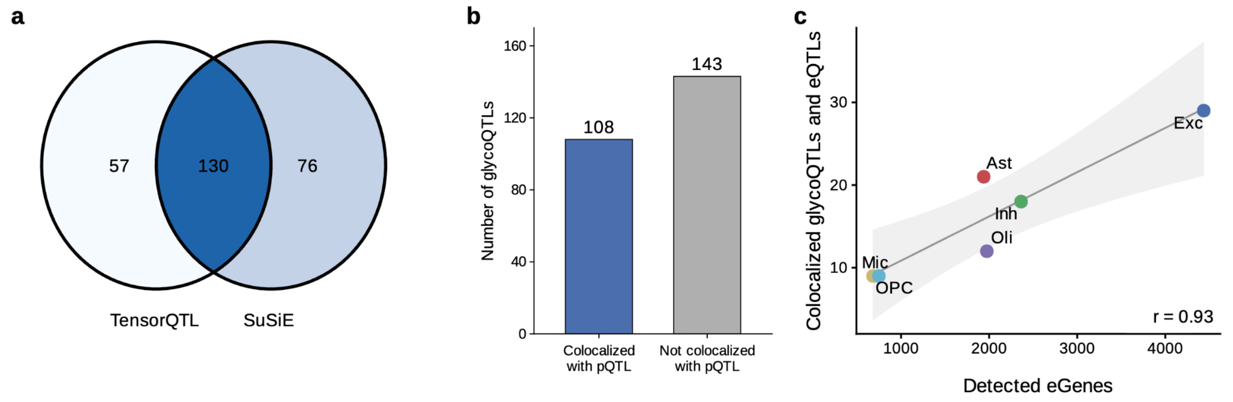


**Supplementary Figure 2. Additional characterization of glycoQTLs.**
a) Overlap between glycopeptiforms with a cis-glycoQTL identified by TensorQTL (permutation test, FDR ≤ 0.05) and SuSiE fine-mapping (95% credible sets) using the unadjusted glycoproteomic data.
b) Number of glycoQTLs with and without evidence of colocalization with pQTLs based on ColocBoost.
c) Relationship between the number of shared glycoQTL/eQTL loci and the number of detected eGenes across six major brain cell types. Cell types with more eGenes also exhibit more shared glycoQTL/eQTL loci (Pearson r = 0.93), indicating that colocalization frequency largely reflects eQTL discovery power rather than cell type-specific enrichment.

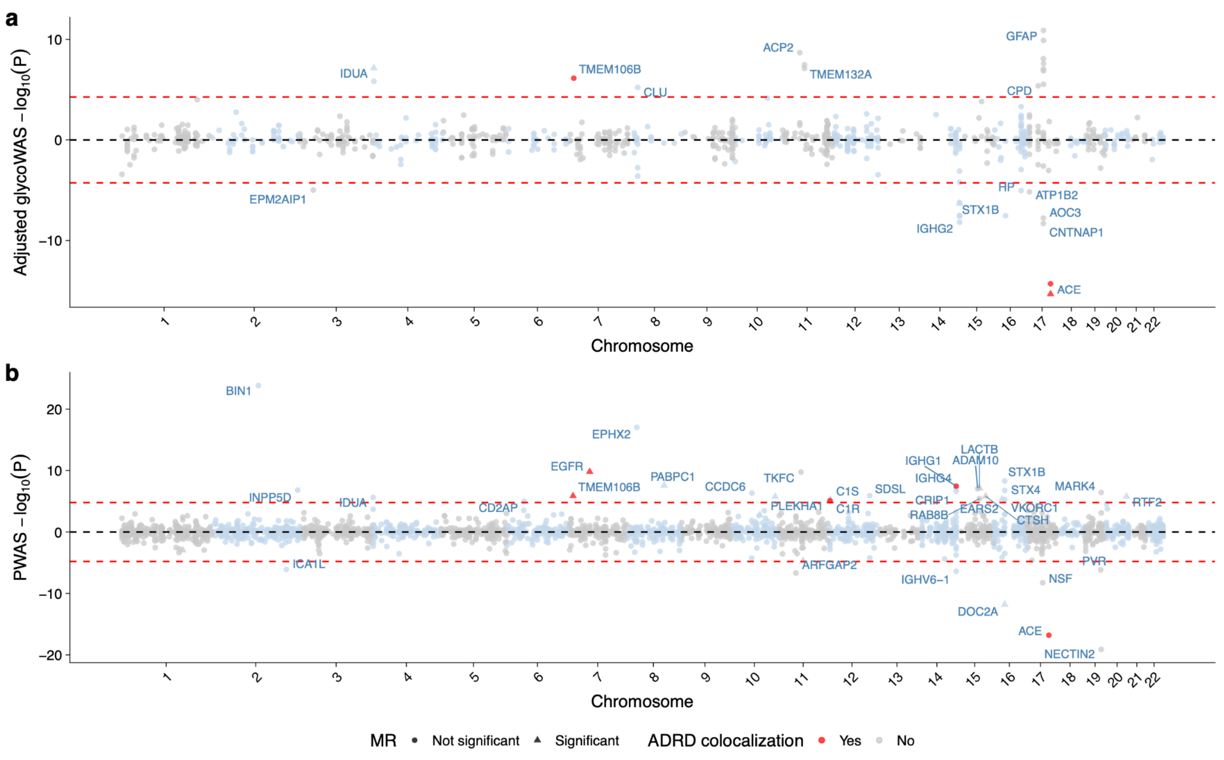


**Supplementary Figure 3. Glycoproteome-wide association and proteome-wide association analyses of Alzheimer's disease and related dementia.**

a) GlycoWAS results using protein-adjusted glycoproteomic data. Each dot represents a glycopeptiform. The association statistic is plotted as the signed log-transformed P value, with positive values indicating that the ADRD risk allele is associated with higher glycopeptiform abundance and negative values indicating lower abundance. Red dashed lines denote the Bonferroni significance threshold. Significant glycopeptiforms are labeled by their corresponding glycoprotein. Among glycopeptiforms with significant GlycoWAS associations, red points indicate glycoQTL colocalization with an ADRD GWAS signal, light blue or gray points indicate no evidence of colocalization, and triangles indicate additional support from Mendelian randomization analysis.

b) PWAS results using matched proteomic data. Each dot represents a protein. The association statistic is plotted as the signed log-transformed P value, with positive values indicating that the ADRD risk allele is associated with higher protein abundance and negative values indicating lower abundance. Red dashed lines denote the Bonferroni significance threshold. Significant proteins are labeled. Among proteins with significant PWAS associations, red points indicate pQTL colocalization with an ADRD GWAS signal, light blue or gray points indicate no evidence of colocalization, and triangles indicate additional support from Mendelian randomization analysis.

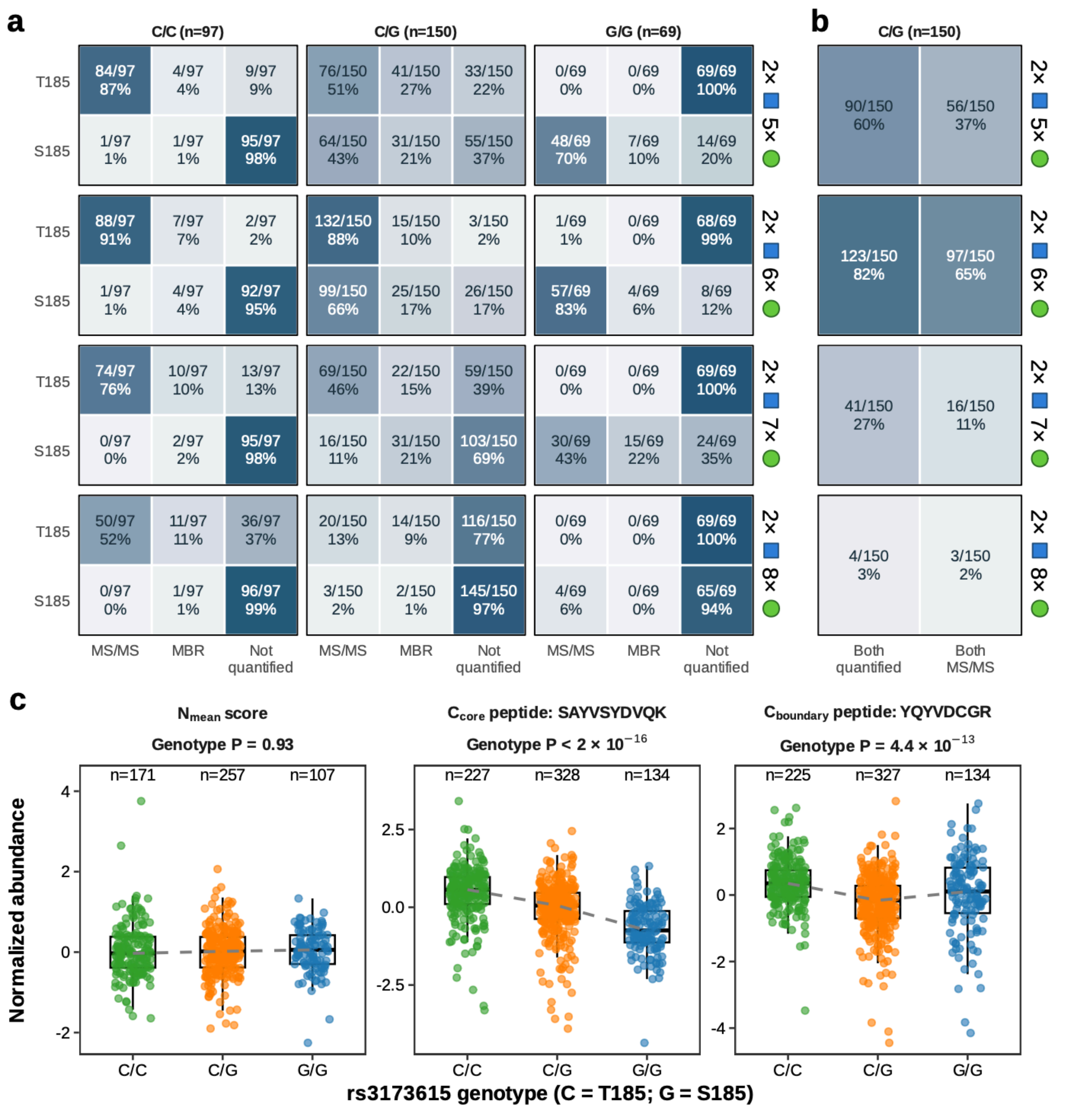


**Supplementary Figure 4. Allele-specific TMEM106B glycopeptiform quantification and tryptic peptide QTLs for rs3173615.**
a) Quantification of T185- and S185-containing glycopeptiforms carrying HexNAc(2)Hex(5-8) at N183, stratified by rs3173615 genotype. Cells show donor counts and percentages for quantification supported by direct, site-localized MS/MS identification (MS/MS), match-between-runs transfer (MBR), or no quantified signal. C encodes the risk T185 allele; G encodes the protective S185 allele. HexNAc(2)Hex(8) was excluded from QTL analyses because of low quantification rates in S185 carriers.
b) Paired-allele quantification in the 150 C/G heterozygotes. For each glycan composition, cells show the number and percentage of donors with both allelic glycopeptiforms quantified and the subset with direct MS/MS support for both.
c) Covariate-adjusted $N_{mean}$ score, $C_{core}$ peptide abundance, and $C_{boundary}$ peptide abundance

by rs3173615 genotype in the TMT cohort. Signals were adjusted for batch, study, age, and sex. $N_{mean}$ combines two standardized N-terminal peptides ($N_1$ and $N_4$).

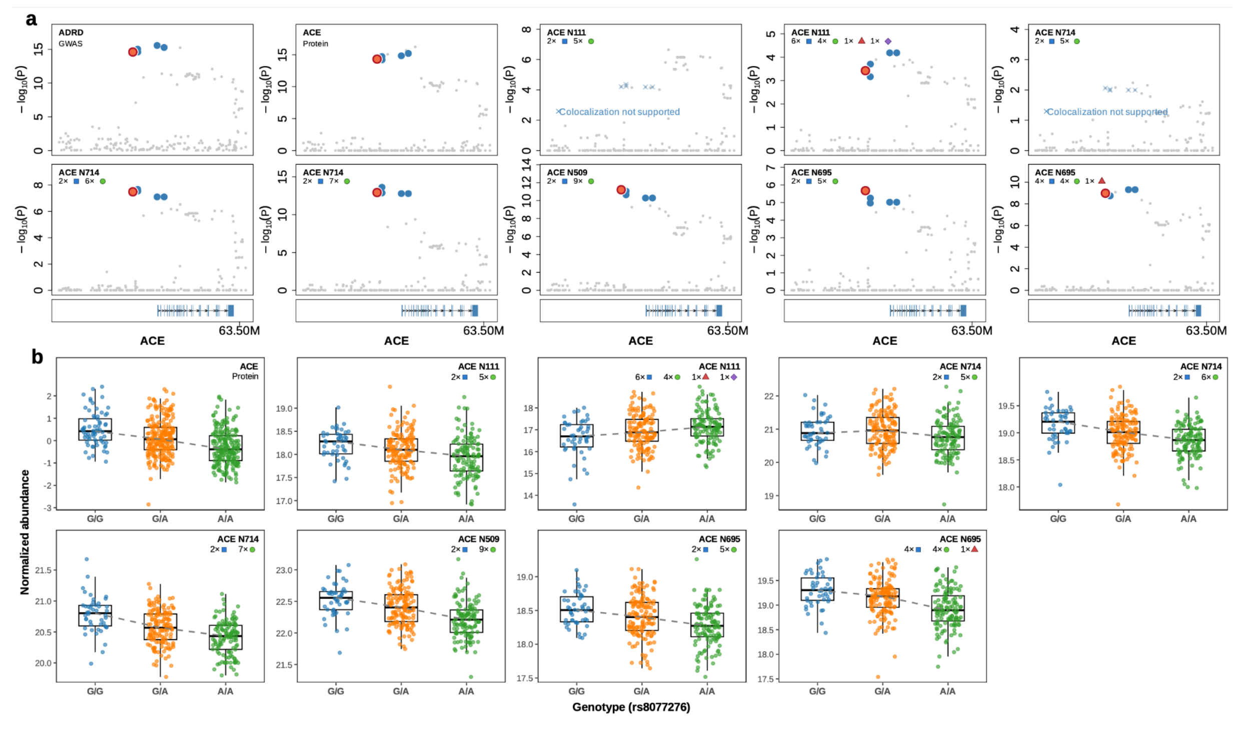


**Supplementary Figure 5. Colocalization and genetic effects of ACE protein and glycopeptiform QTLs.**

a) Association plots for the AD GWAS signal, ACE protein abundance (pQTL), and all eight detected ACE glycopeptiforms at the ACE locus. Each dot represents a genetic variant, with the strength of association shown as the log-transformed P value. Variants in the 95% colocalization confidence set are shown in blue. The lead variant (rs8077276) is highlighted in coral with a red outline.

b) ACE protein abundance and all eight detected ACE glycopeptiforms stratified by rs8077276 genotype. Boxplots show the median and interquartile range. The dashed line connects genotype-group medians.

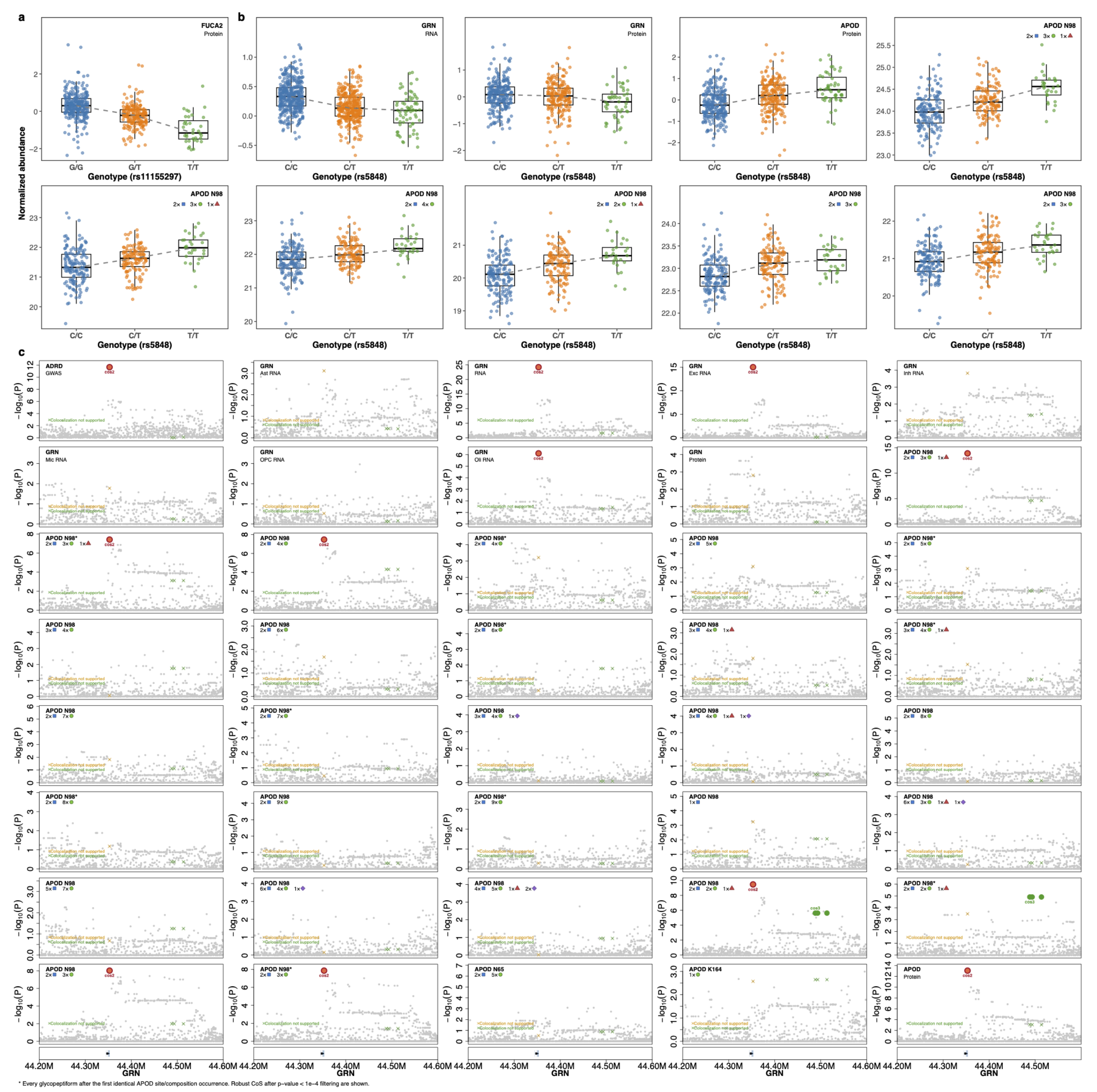


**Supplementary Figure 6. Colocalization across *cis*-GRN and *trans*-APOD glycopeptiforms.**

a) FUCA2 protein abundance stratified by genotype at rs11155297.

b) Abundances of GRN RNA, GRN and APOD proteins and six APOD N98 glycopeptiforms stratified by rs5848 genotype. Boxplots show the median and interquartile range. The dashed line connects genotype-group medians.

c) Colocalization results at the GRN locus for 40 traits tested by ColocBoost: GRN cis-eQTLs across nine brain contexts (Ast, DLPFC, Exc, Inh, Mic, OPC, Oli), the GRN cis-pQTL and APOD trans-pQTL, all 30 APOD trans-glycopeptiforms, and the AD GWAS. Each subpanel shows -log$_{10}$ P against genomic position for one trait. The orange point marks the lead variant of the focal (AD) colocalization; colored crosses mark variants assigned to a credible set of a colocalization signal. All colocalization signals detected at the locus are shown, including the two glycopeptiform-only signals that do not colocalize with the AD GWAS.
